# An experimental model of clinical immunity for human malaria

**DOI:** 10.1101/2025.02.04.25321636

**Authors:** Mimi M. Hou, Adam C. Harding, Natalie M. Barber, Prasun Kundu, Florian A. Bach, Jo Salkeld, Yrene Themistocleous, Nicola M. Greenwood, Jee-Sun Cho, Jordan R. Barrett, Fay L. Nugent, Thomas A. Rawlinson, Susanne H. Hodgson, Baktash Khozoee, Dylan J. Mac Lochlainn, Rachel E. Cowan, Ian D. Poulton, Megan Baker, Lucy Kingham, Celia H. Mitton, Abigail Platt, Raquel Lopez Ramon, Fernando Ramos Lopez, Merin Thomas, Katherine Skinner, Doris Quinkert, Dimitra Pipini, Amelia M. Lias, Martino Bardelli, Nick J. Edwards, Francesca R. Donnellan, Sumi Biswas, Julian C. Rayner, Carolyn M. Nielsen, Sarah E. Silk, Simon J. Draper, Wiebke Nahrendorf, Philip J. Spence, Angela M. Minassian

**Author notes:** joint authors (Adam C. Harding, Natalie M. Barber). joint senior authors (Philip J. Spence, Angela M. Minassian).

## Abstract

Clinical immunity to malaria can reduce fever and lead to asymptomatic infection but the underlying mechanisms remain unclear. To examine the development of clinical immunity, we conducted a multi-cohort, repeat controlled human malaria infection (CHMI) study with *Plasmodium vivax*, and a heterologous rechallenge with *P. falciparum*. Malaria-naïve adults underwent *P. vivax* CHMI up to three times, at an interval of 5 to 20 months, by administration of red blood cells infected with the *P. vivax* PvW1 clone. In the final cohort of the study, a subset of participants underwent heterologous repeat CHMI with the *P. falciparum* 3D7 clone. Clinical parameters and the host response to infection were measured up to 3 months after each CHMI. Nineteen participants underwent primary CHMI with *P. vivax*, 12 returned for secondary homologous CHMI and 2 for tertiary homologous CHMI with the same parasite clone. During rechallenge, parasite growth was not attenuated and there was minimal induction of invasion-blocking antibodies. Nonetheless, clinical symptoms including fever and laboratory abnormalities were less frequent and of lower severity during rechallenge and multi-analyte plasma profiling revealed an attenuated inflammatory response. Six participants who had completed *P. vivax* CHMI, then underwent heterologous rechallenge with *P. falciparum*. Previous infection with *P. vivax* did not protect participants against symptoms, fever or inflammation upon exposure to *P. falciparum*. Clinical immunity to *P. vivax* developed rapidly after a single CHMI, protecting participants against fever and laboratory abnormalities associated with malaria and was underpinned by the attenuation of inflammation. In contrast, there was no evidence of anti-parasite immunity, suggesting that mechanisms of clinical immunity can operate independently of pathogen load to reduce the damage caused by malaria. Clinical immunity to *P. vivax* was parasite species-specific and provided no protection against CHMI with *P. falciparum*.

## Introduction

Malaria remains a major global health burden with an estimated 263 million cases worldwide in 2023^1^. Of the six parasite species that cause human malaria, *Plasmodium falciparum* is the species most commonly associated with severe life-threatening disease and occurs mainly in children under the age of 5 years living in sub-Saharan Africa. *Plasmodium vivax* is often co-endemic with *P. falciparum* and is responsible for more than half of all malaria episodes in the Americas and Southeast Asia^2^. The majority of individuals in West and Central Africa are Duffy-negative and therefore protected from severe *P. vivax* infections. However *P. vivax* can establish low-density infections in Duffy-negative hosts and there is increasing evidence that it is found across the African continent^3,4^. The clinical symptoms of malaria such as fever, headache and fatigue coincide with the onset of systemic inflammation, which leads to abnormalities in clinical laboratory parameters including lymphopaenia and thrombocytopaenia^5,6^. Over a 25-year period, almost half of children living in a high transmission setting in Senegal experienced more than 50 febrile attacks during childhood^7^, which highlights the high burden of malaria on human health. At a population level, malaria is frequently the leading cause of disability-adjusted life years^8^. In the absence of an effective route to local elimination, especially in moderate-to-high transmission settings, malaria control programmes need to prioritise reducing the burden of disease.

Natural exposure to malaria generates clinical immunity, which reduces fever and promotes asymptomatic infection. There are two possible routes to clinical immunity. The first requires parasite control to keep the pathogen load below the pyrogenic threshold^9^. In endemic areas, this takes many years of exposure because it is dependent upon the evolution of an antibody response with sufficient breadth^10,11^. The second is via host adaptations that raise the pyrogenic threshold, which can provide clinical immunity even in the absence of parasite control^12,13^. For *P. falciparum*, this is thought to be a relatively slow process and in agreement with this, we previously found that the first three infections of life trigger similar levels of inflammatory response and symptoms of malaria in a controlled human malaria infection (CHMI) model^14^. In contrast, evidence from endemic areas indicate that the pyrogenic threshold can be raised much more quickly by *P. vivax* infection^15^. Retrospective analyses of records of malaria therapy for the treatment of neurosyphilis patients have shown that clinical immunity can be induced by a single *P. vivax* infection that can lead to strain-transcending protection from fever^16^. However, these historical studies are confounded by the presence of the causative agent of syphilis *Treponema pallidum*, which progressively suppresses the immune system to sustain its own survival^17^.

There are important gaps in our knowledge of how immunity to malaria develops, which may hamper efforts to control this disease. It remains unclear how quickly clinical immunity can develop and how long it can last. We also have no mechanistic understanding of how the pyrogenic threshold can be raised to provide clinical immunity in the absence of parasite control. And we have not examined the interactions between *P. falciparum* and *P. vivax* in an experimental setting. To bridge these gaps, we developed a repeat human malaria challenge model to study the development of clinical immunity to *P. vivax* in detail for the first time in an experimental medicine setting.

## Results

### Study design and participants

In total, 19 malaria-naïve, Duffy blood group positive, adult participants were enrolled and underwent primary blood-stage CHMI with *P. vivax* (**Fig. 1**). Out of these, 12 completed secondary homologous CHMI and 2 completed tertiary homologous CHMI. Following review of results from primary and secondary *P. vivax* CHMI observed in VAC069A-D, the study was amended to study heterologous repeat CHMI with *P. falciparum*. During VAC069E 6 participants completed heterologous *P. falciparum* CHMI: of these, 3 had completed primary CHMI and 3 had completed secondary CHMI with *P. vivax* during VAC069D. Participants were followed up to three months after each CHMI.

**Figure 1.**
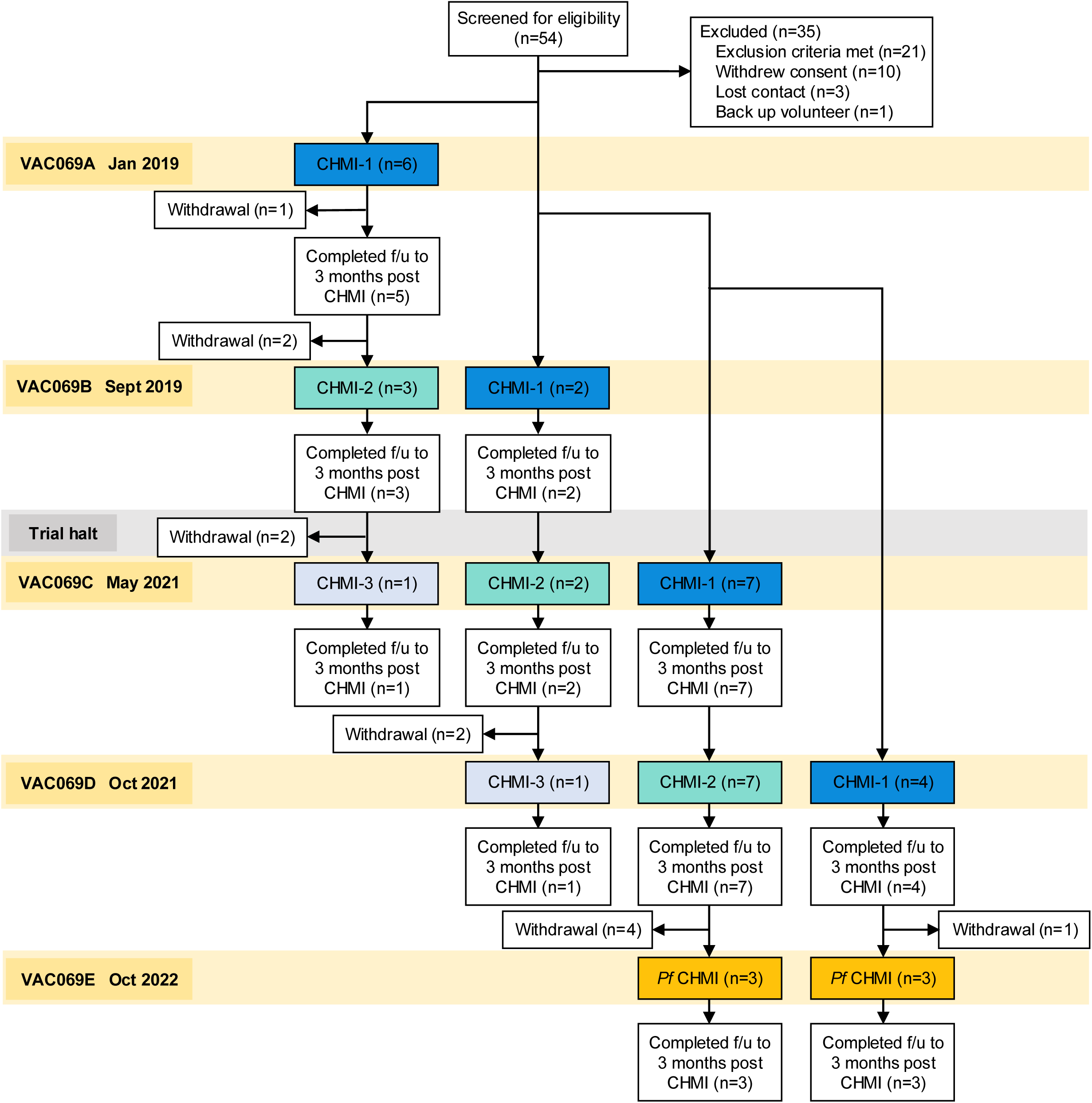
Flow chart of VAC069 study design and participant recruitment. VAC069 was a multicohort study with each cohort (A-E) corresponding to a CHMI. In VAC069A to D, new participants were enrolled to undergo primary CHMI with *P. vivax* (CHMI-1) and participants who had completed CHMI in the previous cohort were invited to undergo secondary homologous CHMI (CHMI-2) followed by tertiary homologous CHMI (CHMI-3). In VAC069E participants who had previously completed one or two CHMIs with *P. vivax* were invited to undergo heterologous CHMI with *P. falciparum* (*Pf* CHMI). Follow-up (f/u) continued to 3 months after each CHMI. The trial was halted during 2020 due to the Covid-19 pandemic.

The VAC079 study, which was conducted alongside VAC069, enrolled 16 participants to test the efficacy of a protein/adjuvant vaccine targeting *P. vivax* Duffy-binding protein region II (PvDBPII) by CHMI. A total of 10 participants completed three vaccinations and underwent primary *P. vivax* CHMI 2 to 4 weeks after their third vaccination, in parallel with unvaccinated participants in VAC069C (**Fig. S1A**). Five participants who completed primary CHMI received a fourth vaccination followed by secondary homologous *P. vivax* CHMI 5 months later, in parallel with participants in VAC069D. Safety, immunogenicity and efficacy data from primary CHMI have previously been published^18^. Here we include the clinical data from primary and secondary CHMI to support the findings of the VAC069 study.

Demographics and Duffy blood group phenotypes of the participants in the VAC069 and VAC079 studies are shown in **Table S1**. These were comparable across all participants undergoing CHMI, except for a preponderance of females in the VAC079 study. For participants who had Duffy blood group phenotypes Fy(a+b-) or Fy(a-b+), their Duffy genotype was determined. In the VAC069 study, one participant with Duffy phenotype Fy(a+b-) and one with phenotype Fy(a-b+) had an erythroid silent FY*B^ES^ allele (Duffy genotype FY*A/FY*B^ES^ and FY*B/FY*B^ES^ respectively), which ablates expression of the Fyb antigen in red blood cells and results in half the level of Fy expression^29^.

No serious adverse events (SAEs) deemed related to CHMI occurred in either the VAC069 or VAC079 studies. All participants in both studies remained seronegative for red blood cell alloantibodies post CHMI and no seroconversion for HIV, hepatitis B and C, Epstein-Barr Virus (EBV) or Cytomegalovirus (CMV) from baseline was observed.

### Parasite multiplication rate is comparable between primary and secondary CHMI

We first asked whether homologous repeat CHMI with the PvW1 clone of *P. vivax* would lead to a reduction in parasite multiplication rate (PMR). All participants developed blood-stage parasitaemia during primary and secondary CHMI (**Fig. 2A, Table S4**). The time taken to reach the protocol-specified malaria diagnosis threshold and the peak parasitaemia were similar between first, second and third infection (**Fig. 2B, S2A-B**). There was no significant difference in PMR between primary CHMI (median PMR = 6.4-fold increase per 48 hours [range 4.0 to 11.1]) and secondary CHMI (median PMR = 6.0 [range 3.8 to 8.4]) (**Fig. 2C**) and PMR was similar in the two participants who underwent tertiary CHMI (8.0 and 8.7) (**S2A**). PMR during primary CHMI was similar in individuals with different Duffy blood group phenotypes, although the two participants with an FY*B^ES^ allele had below average values (**Fig. S2C**). There was no obvious difference in PMR by ethnicity but conclusions are limited by the small number of participants of non-white ethnicity (**Fig. S2D**). In summary, *P. vivax* CHMI does not generate anti-parasite immunity to slow or reduce parasite growth upon rechallenge. Rechallenge was homologous using the same parasite clone, which removes polymorphism as a possible block to the development of immunity. These results reinforce the notion that anti-parasite immunity is acquired slowly and only after many years of repeated exposure to malaria.

**Figure 2.**
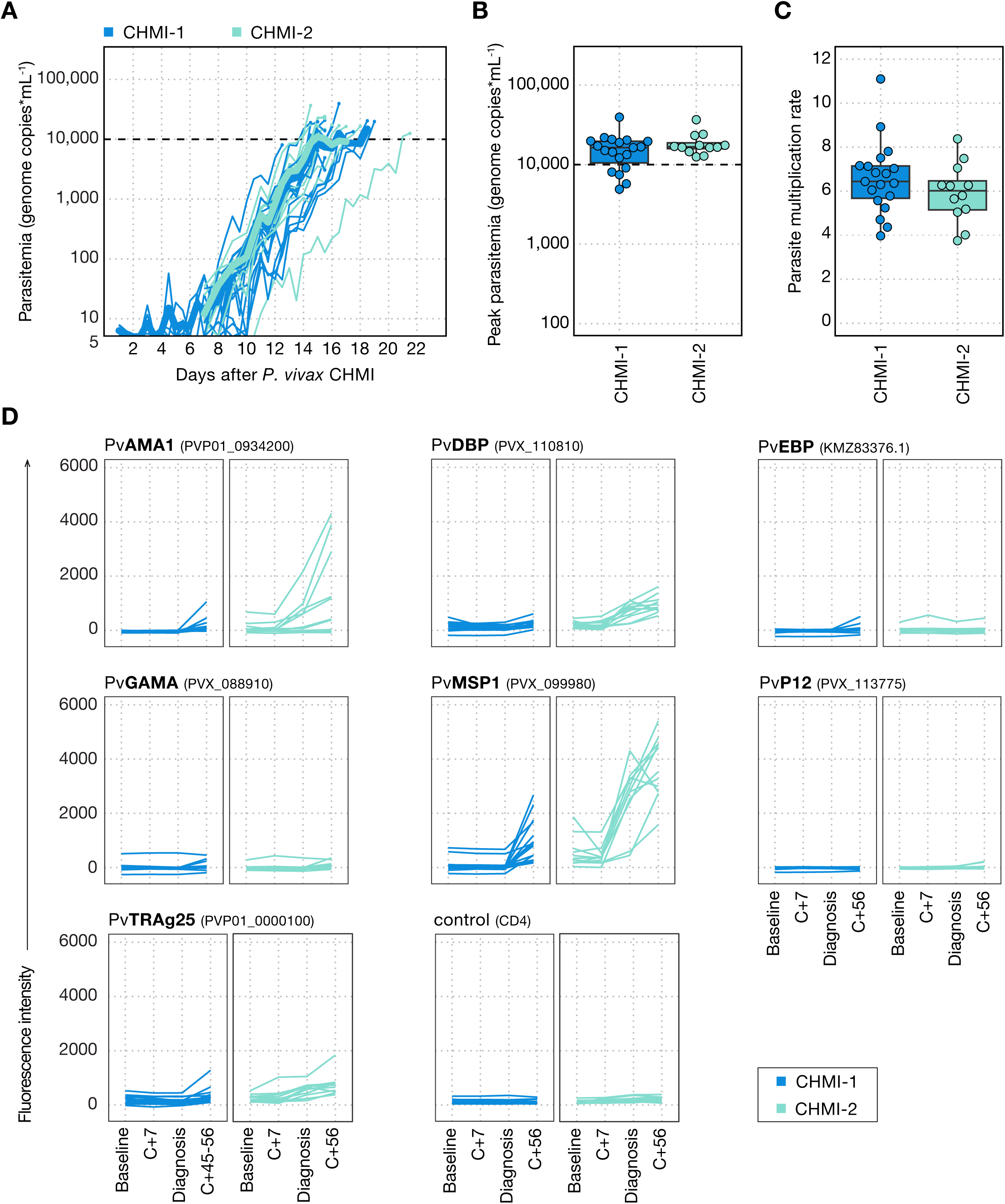
The parasite multiplication rate is comparable between primary and secondary *P. vivax* CHMI despite boosting of anti-merozoite antibodies. **(A)** Parasitaemia was measured up to twice daily by qPCR and is shown for each participant during primary *P. vivax* (CHMI-1) and secondary *P. vivax* CHMI (CHMI-2). The mean parasitaemia is shown in bold and the dashed line indicates the treatment threshold of 10,000 genome copies (gc) ml^-1^. **(B)** Peak parasitaemia for each participant as measured by qPCR during primary and secondary *P. vivax* CHMI. There was no significant difference between paired samples from primary and secondary infection (*p* = 0.7, Wilcoxon signed-rank test) **(C)** Parasite multiplication rate (fold-change per 48 hours) was modelled from each participant’s log_10_ transformed qPCR data. No significant difference was observed between primary and secondary *P. vivax* CHMI (*p* = 0.2, Wilcoxon signed-rank test). **(D)** Serum IgG responses were measured to seven *P. vivax* merozoite antigens using a multiplexed assay: Apical Membrane Antigen 1 (PvAMA1), Duffy Binding Protein (PvDBP), Erythrocyte Binding Protein (PvEBP), GPI-Anchored Micronemal Antigen (PvGAMA), Merozoite Surface Protein 1 (PvMSP1), 6-Cysteine Protein p12 (PvP12) and Tryptophan Rich Antigen 25 (PvTRAg25). Gene IDs are shown in brackets. Antibody responses to CD4 were measured as a negative control. The following timepoints are shown: baseline (1 or 2 days before *P. vivax* challenge), 7 days after challenge (C+7), day of diagnosis and 45 to 56 days after challenge (C+56). In (A-D) n = 19 (primary CHMI) and n = 12 (secondary CHMI).

### Rechallenge leads to boosting of class-switched anti-merozoite antibodies

Anti-parasite immunity is thought to be underpinned by antibody recognition of blood-stage parasite antigens^30^, and so we assessed whether the comparable rates of parasite growth during primary and secondary *P. vivax* CHMI were due to a failure to generate class-switched antibody responses. We used a multiplex immunoassay to measure serum IgG against seven *P. vivax* merozoite antigens: Apical Membrane Antigen 1 (PvAMA1), Duffy Binding Protein (PvDBP), Erythrocyte Binding Protein (PvEBP), GPI-Anchored Micronemal Antigen (PvGAMA), Merozoite Surface Protein 1 (PvMSP1), 6-Cysteine Protein p12 (PvP12) and Tryptophan Rich Antigen 25 (PvTRAg25) (**Fig. 2D**). Antibodies specific for MSP1 were detectable by 56 days after primary CHMI but responses against other merozoite antigens were either very low in a small number of participants or absent. Antibodies against PvMSP1, PvAMA1, PvDBP and PvTRAg25 were all boosted upon rechallenge and already evident at diagnosis. Circulating antibody titres therefore increased during secondary homologous CHMI but, given the comparable parasite growth kinetics during primary and repeat CHMI, these antibodies were evidently insufficient to slow parasite growth.

Antibodies targeting region II of PvDBP have the potential to block merozoite invasion by interrupting the crucial interaction between *P. vivax* and the Duffy antigen expressed on reticulocytes. PvDBPII is the leading blood-stage candidate vaccine antigen and we previously reported in the VAC079 trial that vaccine-induced responses of 150-340 μg/mL, as measured by ELISA, could reduce *P. vivax* growth by ∼50% following primary PvW1 blood-stage^18^. Given we observed responses to the large 140 kDa full-length PvDBP ectodomain in the multiplex immunoassay after secondary CHMI, we assessed whether IgG antibodies specific for region II of PvDBP were also generated during CHMI. However, IgG levels remained undetectable (<1 μg/mL) by ELISA 56 days after primary and secondary CHMI (**Fig. S2E**). Class-switched antibodies specific for merozoite antigens are thus generated and boosted upon a single rechallenge but lack the required breadth and/or specificity to effectively neutralise reticulocyte invasion.

### A single infection induces long-lived mechanisms of clinical immunity

Clinical immunity can be generated even in the absence of parasite control^12,13^ and has been observed after one malaria infection in neurosyphilis patients^16^. We therefore assessed whether the frequency or severity of symptoms was altered between first and second CHMI. In VAC069 at least one solicited adverse event (AE) was reported by all participants during both primary and secondary CHMI. During primary CHMI, solicited AEs increased in frequency and severity around the time of diagnosis, peaked within 48 hours and mostly resolved by six days after starting drug treatment (**Fig. S3A**). A minority of participants reported AEs relating to anti-malarial drugs, which resolved quickly upon completion of treatment (**Fig. S3B**). Unsolicited AEs assessed to be at least possibly related to primary CHMI were predominantly of mild severity, with decreased appetite the most frequent symptom reported (**Table S2A**). The maximum severity of any solicited AE was markedly reduced during rechallenge, with only 1 out of 12 (8%) participants undergoing secondary CHMI reporting at least one severe (grade 3) AE, compared to 9 out of 19 (47%) participants undergoing primary CHMI (**Fig. 3A**). The most commonly reported solicited AEs were headache, fatigue and malaise. All solicited AEs occurred less frequently and with lower severity during secondary compared to primary CHMI (**Fig. 3B**).

**Figure 3.**
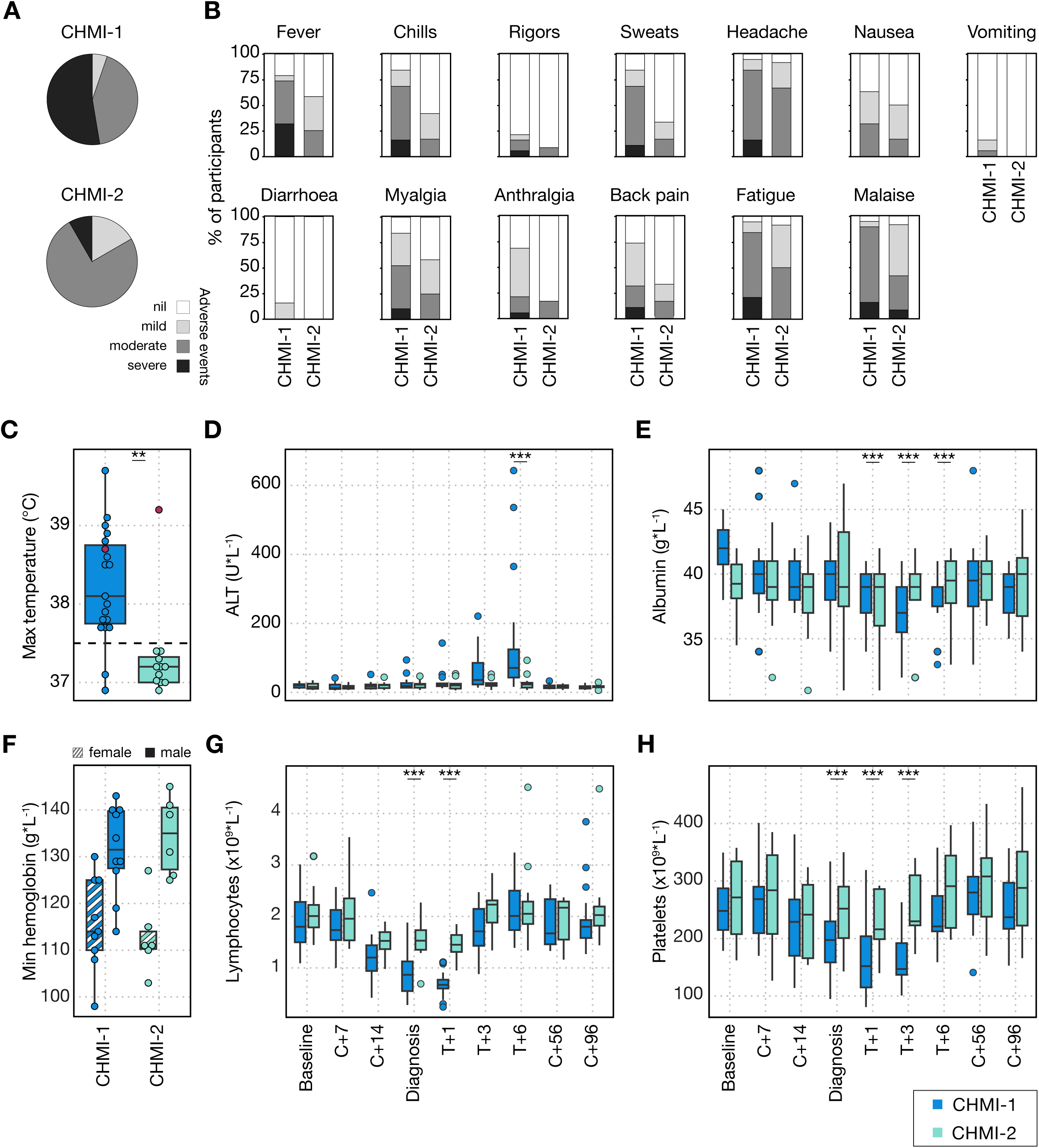
A single *P. vivax* infection induces long-lived mechanisms of clinical immunity. (**A-B**) Clinical signs and symptoms of malaria in participants undergoing primary *P. vivax* CHMI (CHMI-1) compared to secondary *P. vivax* CHMI (CHMI-2) in the VAC069 study. Data are shown as a proportion of the total number of participants undergoing CHMI. **(A)** shows the maximum severity of any solicited adverse event (AE) reported by an individual in the 48 hours before and after diagnosis; **(B)** shows the frequency and severity of each solicited AE. **(C)** Maximum recorded core temperature during primary and secondary *P. vivax* CHMI (*p* < 0.01 by Wilcoxon signed-rank test for paired samples). The pink dots represent the one participant who experienced fever upon rechallenge. **(D-H)** Laboratory parameters (biochemistry and full blood counts) were measured during primary and secondary *P. vivax* CHMI at baseline (1 or 2 days before challenge); 7 and 14 days after challenge (C+7 and C+14); on the day of diagnosis; 1, 3 and 6 days after treatment (T+1, T+3 and T+6); 45 to 56 days (C+56) and 96 days (C+96) after challenge. Statistically significant differences between CHMI-1 and CHMI-2 were identified at each time-point by using mixed-effects modelling and linear regression (*** *p* < 0.001). **(D)** shows alanine aminotransferase (ALT); **(E)** shows albumin. **(F)** shows the minimum haemoglobin concentration with data split by participant sex. There was no significant difference between CHMI-1 and CHMI-2 (Wilcoxon signed-rank test, *p* = 0.8 for females and *p* = 0.9 for males). **(G)** shows lymphocyte count; **(H)** shows platelet count. Box and whisker plots show median and interquartile range (IQR) with whiskers representing 1.5x IQR (outliers are shown as dots). In (A-F) n = 19 (primary *P. vivax* CHMI) and n = 12 (secondary *P. vivax* CHMI).

Symptoms reported by participants are subjective so we assessed whether there was a comparable reduction in the frequency and severity of quantifiable clinical and laboratory parameters (**Table S3A**). Pyrexia (temperature >37.5°C) was recorded in 17 out of 19 (89%) participants during primary CHMI but in only 1 out of 12 participants (8%) upon rechallenge. Consequently, the maximum recorded temperature was significantly lower in secondary CHMI (median = 37.2°C) compared to the same 12 participants during primary CHMI (median = 38.3°C, Wilcoxon signed rank test *p* < 0.01) (**Fig. 3C**). Raised alanine aminotransferase (ALT) (>55 IU/L) was observed in 12 out of 19 (63%) participants undergoing primary CHMI, with ALT levels peaking around six days post-treatment and normalising by 2 months after challenge (**Fig. 3D**). This was severe in three participants with the highest recorded ALT at 14 times the upper limit of normal. All participants with raised ALT had normal alkaline phosphatase (ALP) and bilirubin levels, and clotting tests performed in those with severe transaminitis also remained normal. ALT at day 6 post-treatment was significantly lower during secondary compared to primary CHMI and transaminitis was only observed in 1 out of 12 (8%) participants during secondary CHMI. Of note, this individual continued to have significant malaria symptoms and was the only participant who was febrile upon both secondary and tertiary rechallenge (**Fig. 3C**). Serum albumin was also significantly lower at days 1-6 post-treatment during primary CHMI compared to during rechallenge (**Fig. 3E**).

We observed a slight reduction in haemoglobin 6 days after starting antimalarial treatment but these did not differ between primary and secondary CHMI (**Fig. 3F**). In contrast, lymphopaenia was pronounced, reaching a nadir at 24 hours after diagnosis and was significantly attenuated upon rechallenge (linear regression *p* <0.001) (**Fig. 3G**). Thrombocytopaenia, also lowest at 24 hours after treatment, was similarly significantly attenuated during secondary compared to primary CHMI (linear regression *p* <0.001) (**Fig. 3H**). Taken together, these data indicate that participants are protected against pyrexia, liver injury, lymphopaenia and thrombocytopaenia when undergoing homologous repeat *P. vivax* CHMI. In concordance with these results, we also observed reduced signs and symptoms of malaria after rechallenge in the VAC079 study. In the VAC079 study, the rate of fever and the severity of solicited AEs and laboratory abnormalities such as transaminitis were similar to VAC069 during primary CHMI but were significantly reduced during secondary CHMI (**Table S3B**, **Fig. S1B-D**). Unsolicited AEs, which were mostly mild in severity, were also reported less frequently during secondary compared to primary CHMI (**Table S2B**). A single infection with *P. vivax* is therefore sufficient to generate and maintain mechanisms of long-lived clinical immunity (20 months in this study) that can reduce the harm caused by malaria parasites during subsequent infections and these mechanisms are independent of anti-parasite immunity.

### Clinical immunity is underpinned by attenuated inflammation

Many of the symptoms of malaria such as fever and laboratory abnormalities including lymphopaenia are caused by the host response to infection^31^. We hypothesised that systemic inflammation would be attenuated upon rechallenge to raise the pyrogenic threshold and improve clinical outcome. We therefore measured plasma analytes indicative of inflammation, coagulation, oxidative stress and tissue damage using a bead-based multiplexed protein assay and compared the dynamics of each analyte through time during primary and secondary CHMI. For these experiments, we selected 7 participants who underwent primary CHMI in VAC069C and secondary CHMI in VAC069D, as well as 3 participants who underwent primary CHMI in VAC069D. Samples were not used from VAC069A or VAC069B because fewer samples from post-treatment time-points were available for analysis.

We found that plasma levels of the major pyrogenic cytokines or their regulators interleukin 1 receptor A (IL-1RA), interleukin 6 (IL-6) and soluble tumor necrosis factor receptor 2 (sTNFRII) were raised at diagnosis and peaked 24 hours after drug treatment during primary CHMI (**Fig. 4A**). However, these inflammatory markers were significantly reduced upon rechallenge. The interferon-stimulated C-X-C motif chemokine ligand 10 (CXCL10), the critical host factor for recruitment of T cells into inflamed tissues, as well as the cytokines IL-12p70 and IL-18, which promote T cell activation and differentiation, were also attenuated during rechallenge (**Fig. 4B**). Similarly, markers of coagulation and endothelium activation, which peaked between 1 and 3 days after diagnosis in primary CHMI, were reduced upon rechallenge and remained at almost baseline levels during and after secondary CHMI (**Fig. 4C**). Our data reveal that *P. vivax* can rapidly induce host adaptations that restrict inflammation, avert a pro-coagulant state and prevent endothelial dysfunction to raise the pyrogenic threshold and minimise the clinical symptoms associated with malaria.

**Figure 4.**
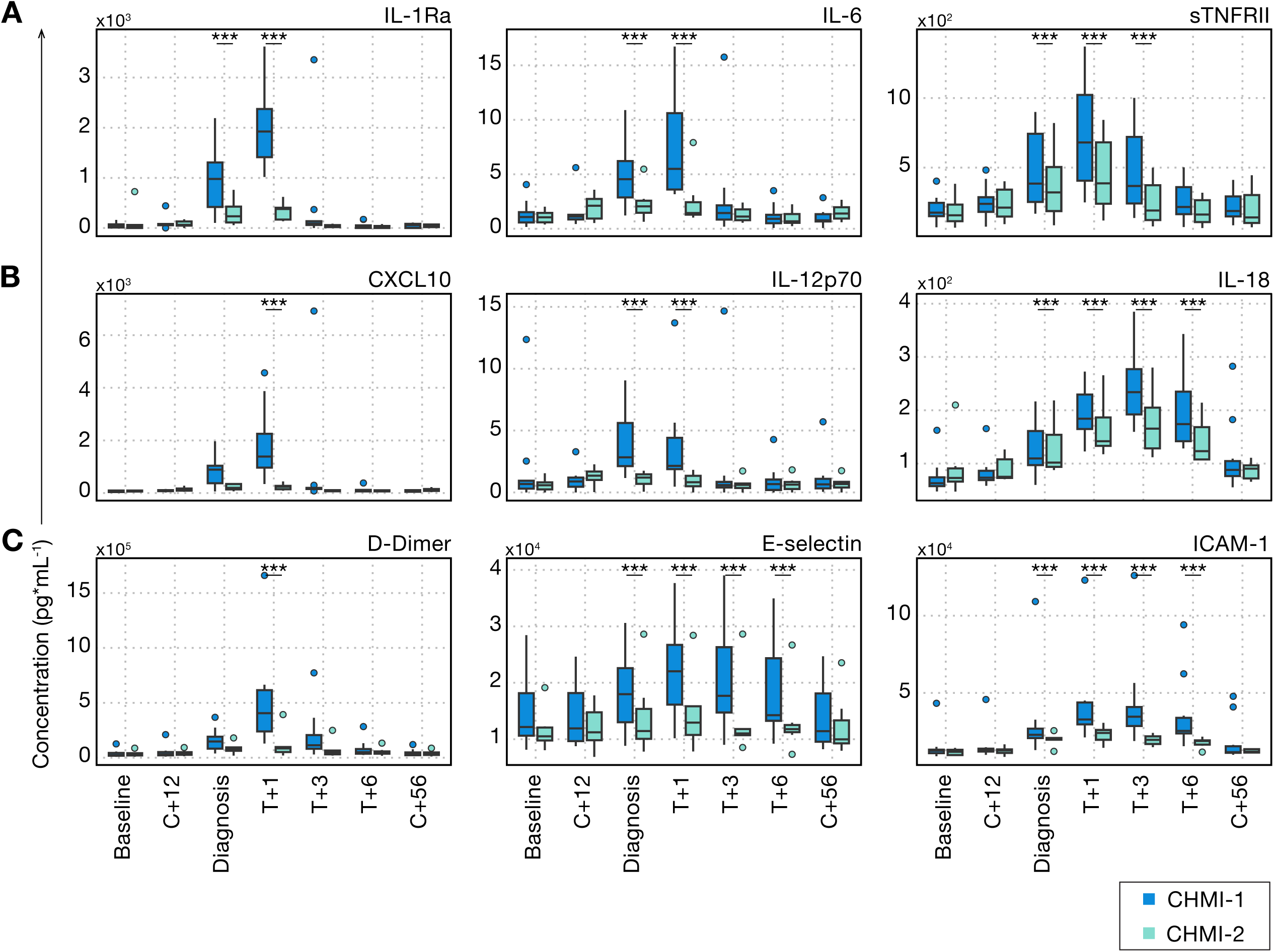
Clinical immunity to *P. vivax* is underpinned by attenuated inflammation. **(A-C)** Circulating biomarkers of inflammation, coagulation and endothelial cell activation were quantified during and after primary and secondary *P. vivax* CHMI using a bead-based multiplexed protein assay. We analysed plasma proteins at baseline (1 or 2 days before challenge); 12 days after challenge (C+12); on the day of diagnosis; 1, 3 and 6 days after treatment (T+1, T+3 and T+6); and 45 to 56 days after challenge (C+56). **(A)** shows pyrogenic cytokines and their regulators, **(B)** shows chemokines and cytokines involved in T cell recruitment and activation, and **(C)** shows markers of coagulation and endothelial cell activation. Box and whisker plots show median and interquartile range (IQR) with whiskers representing 1.5x IQR (outliers are shown as dots). Statistically significant differences between CHMI-1 and CHMI-2 were identified at each time-point by using mixed-effects modelling and linear regression (*** *p* < 0.001). In (A-C) n = 10 (primary *P. vivax* CHMI) and n = 7 (secondary *P. vivax* CHMI).

### Clinical immunity is parasite species-specific

Given that *P. vivax* is co-endemic with *P. falciparum* across much of the world^2–4^, we next wanted to assess whether clinical immunity is specific to the parasite species that raised the pyrogenic threshold. Neurosyphilis patients were sometimes infected with *P. falciparum* once they became refractory to *P. vivax* -induced fever^32^. We therefore hypothesised that clinical immunity which developed against *P. vivax* would not be effective against rechallenge with *P. falciparum*. To test this hypothesis, we amended VAC069E to infect participants who had previously undergone one or two prior *P. vivax* CHMIs, with the 3D7 clone of *P. falciparum* using a blood challenge^21^. *P. falciparum*, which preferentially invades mature red cells, has a higher PMR than *P. vivax*, which is restricted to reticulocytes. In concordance with this, we found that in VAC069E, participants infected with *P. falciparum* reached the treatment threshold quicker than when previously infected with *P. vivax* (**Fig. 5A**). Peak parasitaemia was comparable to primary and secondary *P. vivax* CHMI, therefore any differences between homologous and heterologous rechallenge in the VAC069 study were not confounded by circulating pathogen load. *P. falciparum* growth dynamics and the time to reach malaria diagnostic criteria were also similar to those seen in primary blood-stage *P. falciparum* CHMI in our previous studies VAC054 and VAC063 conducted in Oxford using the same 3D7 clone^21,33,34^ (**Fig. S4A-B**).

**Figure 5.**
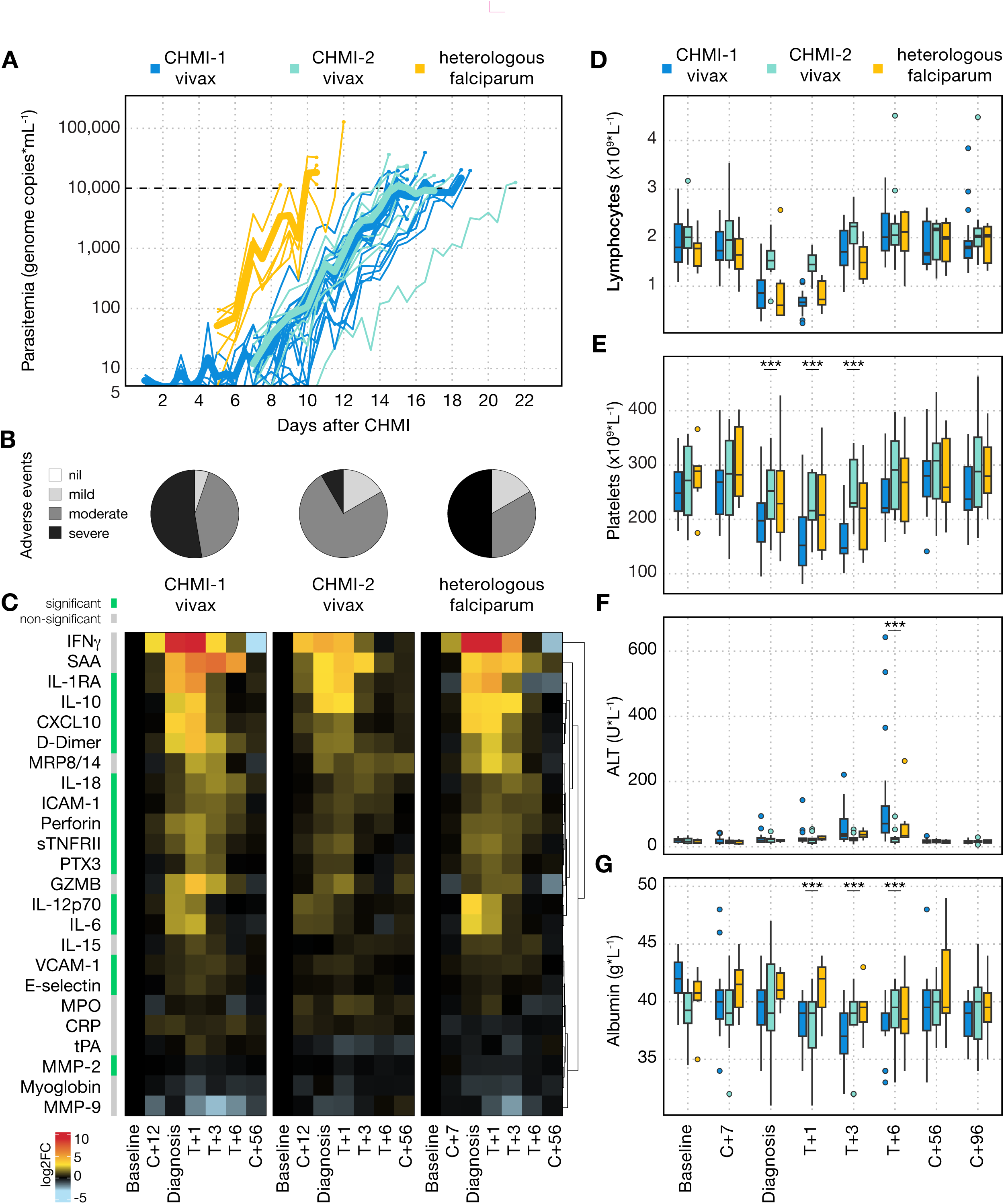
Clinical immunity is parasite species-specific. **(A)** Parasitaemia was measured up to twice daily by qPCR during primary *P. vivax* (CHMI-1) and secondary *P. vivax* CHMI (CHMI-2) as well as during heterologous rechallenge with *P. falciparum*. The same diagnosis and treatment criteria were used during each CHMI. The mean parasitaemia is shown in bold and the dashed line indicates the treatment threshold of 10,000 genome copies (gc) ml^-1^. **(B)** shows the maximum severity of any solicited adverse event (AE) reported by an individual in the 48 hours before and after diagnosis during primary and secondary *P. vivax* CHMI as well as during heterologous *P. falciparum* rechallenge. Data are shown as a proportion of the total number of participants undergoing CHMI. **(C)** Heatmap showing the log_2_ fold-change of 24 plasma analytes during primary and secondary *P. vivax* CHMI as well as during heterologous *P. falciparum* rechallenge. Data are shown relative to baseline values at the following time-points: 7 (C+7) or 12 (C+12) days after challenge with *P. falciparum* or *P. vivax*, respectively; day of diagnosis; 1, 3 and 6 days after treatment (T+1, T+3, T+6); and 45 to 56 days after challenge (C+56). Analytes are ordered by unsupervised hierarchical clustering and those that vary significantly between primary and secondary *P. vivax* CHMI *and* between secondary *P. vivax* CHMI and *P. falciparum* rechallenge are indicated in green in the bar to the left of the heatmap (grey is non-significant). Significance was assessed using mixed-effects modelling and linear regression. The data for each CHMI are paired and for *P. vivax* the raw data for 9 of these analytes are shown in Figure 4. **(D-G)** Laboratory parameters (biochemistry and full blood counts) were measured during primary and secondary *P. vivax* CHMI as well as during heterologous *P. falciparum* rechallenge. Data are shown at baseline (1 or 2 days before challenge); 7 days after challenge (C+7); on the day of diagnosis; 1, 3 and 6 days after treatment (T+1, T+3 and T+6); and 45 to 56 days (C+56) as well as 96 days (C+96) after challenge. Statistically significant differences between primary *P. vivax* CHMI and heterologous *P. falciparum* rechallenge were identified at each time-point using mixed-effects modelling and linear regression (*** *p* < 0.001). **(D)** shows lymphocyte count; **(E)** shows platelet count; **(F)** shows alanine aminotransferase (ALT); **(G)** shows albumin. Box and whisker plots show median and interquartile range (IQR) with whiskers representing 1.5x IQR (outliers are shown as dots). In (A-B) and (D-G) n = 19 (primary *P. vivax* CHMI), n = 12 (secondary *P. vivax* CHMI) and n = 6 (heterologous *P. falciparum* CHMI). In (C) n = 10 (primary *P. vivax* CHMI), n = 7 (secondary *P. vivax* CHMI) and n = 6 (heterologous *P. falciparum* CHMI).

Clinical symptoms during VAC069E occurred at a similar frequency and severity to primary *P. vivax* CHMI (**Fig. 5B, Table S2C, Table S3C**) and were comparable to primary *P. falciparum* infections in our previous studies *P. falciparum* CHIM studies VAC054 and VAC063 (**Fig. S4C**)^21,33,34^. The proportion of participants experiencing fever in VAC069E (2 out of 6 [33%] participants) did not significantly deviate from that observed during primary CHMI in the VAC054 and VAC063 studies (23 out of 39 [59%] participants)^21,33,34^ (Barnards CSM exact test *p = 0.3*). During VAC069E the inflammatory and coagulation markers that were attenuated when participants underwent secondary *P. vivax* CHMI (including IL-6, CXCL10 and D-Dimer) were all significantly increased again during heterologous *P. falciparum* rechallenge (**Fig. 5C**). Heterologous *P. falciparum* rechallenge led to a pronounced lymphopaenia that was comparable to primary *P. vivax* CHMI (**Fig. 5D**), whereas thrombocytopaenia and liver injury remained attenuated during heterologous *P. falciparum* rechallenge compared to primary *P. vivax* CHMI (**Fig. 5E-G**). When compared to primary *P. falciparum* CHMI in the VAC054 and VAC063 studies, lymphopaenia and thrombocytopaenia were comparable in those undergoing heterologous *P. falciparum* rechallenge in VAC069E (**Fig. S4D-E**). In our historical studies VAC054 and VAC063, no blood tests were taken at day 6 post-treatment, at the peak of ALT rise, so we cannot compare rates of transaminitis to primary *P. falciparum* CHMI. Our data show that infection with *P. falciparum* after prior exposure to *P. vivax* leads to systemic inflammation, fever and lymphopenia comparable to that observed during primary *P. vivax* and primary *P. falciparum* CHMI. Clinical immunity as defined by symptomatology is thus parasite species-specific.

## Discussion

CHMI and rechallenge studies provide a unique opportunity to investigate mechanisms of naturally acquired immunity *in vivo*. Here we report the safety and infectivity, as well as the parasite growth dynamics and host response during primary and secondary homologous *P. vivax* CHMI using the PvW1 clone^19^, followed by heterologous CHMI with the *P. falciparum* 3D7 clone. We show that homologous and heterologous rechallenge is safe in malaria naïve adults and all participants were reliably infected after each inoculation.

The PMR during primary and secondary homologous *P. vivax* CHMI was comparable, suggesting participants did not develop effective mechanisms of anti-parasite immunity during the VAC069 study. This is in agreement with our previous CHMI study in which participants showed no change in *P. falciparum* PMR after three infections^33,34^. These findings are surprising as polymorphism, thought to be a major barrier to the development of anti-parasite immunity, is not a concern in homologous rechallenge studies that use clonal parasites as has been done in our studies. These results are also at odds with the findings of studies of malaria therapy patients, which often reported a reduced parasitaemia upon homologous reinfection for both *P. vivax* and *P. falciparum* ^16,35^. This difference in outcome is unlikely to be due to the long interval between each CHMI in our studies (up to 20 months in VAC069) because intervals of up to 96 months were found to have no relationship with the degree of anti-parasite immunity in malaria therapy studies^6^. Instead, we suggest that this difference is best explained by the lower treatment threshold applied during CHMI studies. The maximum parasitaemia in VAC069 was approximately 1000 times lower than that reported in patients undergoing malaria therapy^16^ and 10,000 times lower when compared to a recent *P. vivax* rechallenge study in non-human primates^36^. As such, even though we observe antibody boosting at diagnosis during rechallenge, participants may be drug-treated before the humoral response is boosted significantly enough to impact parasite growth. Alternatively, the short duration of CHMI may generate antibodies that lack the required breadth and/or specificity to effectively neutralise reticulocyte invasion. High concentrations of vaccine-induced antibodies specific for region II of PvDBP were able to slow parasite growth in a CHMI model^18^ but these antibodies were not induced at any significant magnitude by *P. vivax* infection during VAC069.

The absence of anti-parasite immunity in CHMI thus allows us to explore mechanisms of clinical immunity that can operate independently of pathogen load. These are thought to be generated very quickly against *P. vivax* in endemic areas and support the early transition to asymptomatic infection^15^. In agreement, we find that a single CHMI leads to robust and long-lived protection against clinical symptoms, fever and laboratory abnormalities associated with *P. vivax* malaria, which was maintained for up to 20 months before rechallenge. Furthermore, protection coincides with the attenuation of systemic inflammation, coagulation and endothelium activation. We can therefore conclude that the pyrogenic threshold can be raised sufficiently after one infection with *P. vivax* in healthy adults to generate clinical immunity. In contrast, in our previous repeat *P. falciparum* CHMI study, no difference in the rate of fever was observed in those undergoing secondary or tertiary homologous rechallenge^34^.

But what are the mechanisms that can minimise inflammation and reduce harm after a single episode of *P. vivax* malaria? Many of the inflammatory markers we measured in VAC069 are produced by innate immune cells, including monocytes and neutrophils, as well as non-immune cells, such as fibroblasts. These diverse cell-types cannot acquire memory in the same way as adaptive T and B cells and may instead be epigenetically modified to reduce their responsiveness to parasites and their pyrogenic products. Epigenetic reprogramming of innate immune and non-haematopoietic cells has been termed “innate memory”, and has been shown to underpin endotoxin tolerance *in vitro* and *in vivo* ^37^. Furthermore, monocyte reprogramming has been shown to regulate inflammation in human sepsis patients^38^. So does malaria similarly induce innate memory? *In vitro* exposure of human monocytes to parasitised red blood cells leads to epigenetic modifications and a reduced inflammatory response to re-stimulation^39^. Similarly, CHMI modifies the deposition of H3K4me3 at the promoter regions of inflammatory genes, leading to long-term functional changes in monocytes that can persist for several weeks after parasite clearance^40^. More recently, asymptomatic infection in children living in malaria-endemic Uganda was shown to correlate with the epigenetic modification of circulating monocytes^41^. These studies assessed the impact of malaria on short-lived terminally differentiated effectors, whereas long-term memory would require epigenetic reprogramming of progenitor cells in the bone marrow^42^. Future studies that combine CHMI with bone marrow sampling such as our ongoing BIO-004 trial (ISRCTN85988131) will therefore allow us to directly test the hypothesis that innate memory underpins clinical immunity to malaria. We will also be able to investigate alternative hypotheses such as malaria-induced remodelling of the tissue microenvironment to regulate innate immune cell function^43^.

To date, studies of innate memory have mainly focussed on *P. falciparum* and this raises two important questions surrounding our CHMI studies. If we observe clinical immunity after a single *P. vivax* episode, why was this not induced in participants who were enrolled in our repeat blood-stage *P. falciparum* CHMI trial^33,34^? And why is clinical immunity species-specific, when the parasite products that trigger inflammation and pyrexia, such as DNA and hemozoin, are shared by all parasite species? Our results are in agreement with observations from endemic areas^15^ as well as retrospective analyses of malaria therapy records^16,35,44^ so our observations are unlikely to be an artefact of CHMI. Instead, the answer to both of these questions may relate to differences in parasite DNA motifs recognised by the innate immune system. For example, *P. vivax* preferentially triggers TLR9 via CpG motifs whereas the *P. falciparum* genome is more AT-rich and preferentially triggering TLR9-independent pathways^45^. These biological differences may be further compounded by the preference of *P. vivax* for the bone marrow parenchyma^46^, which is where the innate immune response to blood-stage infection is thought to be primed^47^. We therefore need to better understand how differences in parasite biology influence innate immune cells in the short- and long-term, and heterologous CHMI is an effective way to test clinical immunity against different *Plasmodium* species. It is notable that *P. malariae* may protect against *P. falciparum* ^44^, and the development of a *P. malariae* blood bank for CHMI^48^ offers a tantalising opportunity to gain further mechanistic insight into clinical immunity.

We remain short of having all of the tools required to pursue widespread eradication of *P. falciparum* and *P. vivax,* and so disease control programmes will likely be at the forefront of public health policy for years to come. Here, we show that a single infection with *P. vivax* induces long-lasting clinical immunity that can prevent fever and symptoms of malaria in the absence of effective anti-parasite immunity. This lays the groundwork for a mechanistic understanding of clinical immunity to malaria in people and could accelerate the development of interventions that specifically reduce the clinical severity and morbidity of malaria disease.

## Methods

### Study design and participants

VAC069 was a multi-cohort study to evaluate the safety and feasibility of repeat blood-stage *P. vivax* CHMI. During each of the five cohorts of the study VAC069A-E, participants underwent CHMI in parallel. Malaria-naïve, Duffy blood group positive adults aged 18 to 50 years were enrolled to the study at the Centre for Clinical Vaccinology and Tropical Medicine, University of Oxford, UK. Full inclusion and exclusion criteria are provided in the Supplementary Appendix. Participants who completed primary CHMI with *P. vivax* were invited to undergo secondary, followed by tertiary homologous CHMI during subsequent cohorts of the study. New participants were enrolled to undergo primary CHMI in parallel with participants undergoing repeat CHMI. Repeat CHMI took place at intervals of 5, 8 and 20 months; the 20 months interval was due to the study being temporarily halted during the COVID-19 pandemic. In VAC069E, participants who had previously completed primary or secondary CHMI with *P. vivax* were invited to undergo heterologous CHMI with *P. falciparum* (**Fig. 1**).

Data from all participants undergoing primary CHMI with *P. vivax* across four cohorts VAC069A–D were pooled (n = 19) and compared to pooled data from all participants undergoing secondary homologous CHMI across three cohorts VAC069B–D (n = 12). All 6 participants who received heterologous *P. falciparum* CHMI during VAC069E were pooled for analysis.

VAC079 was a phase I/IIa trial to assess the efficacy of the protein vaccine PvDBPII in Matrix-M adjuvant by CHMI. Participants received three doses of the vaccine, followed by primary CHMI with *P. vivax*, previously reported elsewhere^18^. A subset of participants received a fourth vaccination and underwent secondary homologous CHMI 5 months after their primary CHMI. Participants from the VAC079 study underwent *P. vivax* CHMI in parallel with participants in the VAC069 study.

### Trial oversight

VAC069 and VAC079 were designed and conducted at the University of Oxford. The studies are registered on ClinicalTrials.gov (VAC069: NCT03797989; VAC079: NCT04201431) and received ethical approval from UK National Health Service Research Ethics Services (VAC069: Hampshire A Research Ethics Committee, Ref 18/SC/0577; VAC079: Oxford A Research Ethics Committee, Ref 19/SC/0330). VAC079 was approved by the UK Medicines and Healthcare products Regulatory Agency (EudraCT 2019-002872-14). All participants provided written consent. The studies were conducted according to the principles of the current revision of the Declaration of Helsinki 2008 and ICH guidelines for Good Clinical Practice.

### Blood-stage Controlled Human Malaria Infection

*P. vivax* CHMI was initiated by intravenous injection of red blood cells infected with the PvW1 clone, which originates from Thailand. This cryopreserved *P. vivax* blood stabilate was produced from a blood group O-donor at the University of Oxford who was infected by mosquito bite^19^ since mosquitoes have been shown to reset *Plasmodium* virulence^20^. On the day of CHMI, thawed aliquots of cryopreserved PvW1 inoculum were made up to 5 mL in normal saline and injected intravenously^19,21^. In VAC069A 2 participants each received one vial of inoculum, 2 participants each received one fifth of a vial and 2 participants each received one twentieth of a vial. In subsequent CHMIs in VAC069B-D and the VAC079 study, all participants received one tenth of a vial of inoculum during each CHMI. One vial of inoculum contained between 1650 to 2170 genome copies (gc) of *P. vivax* as measured by quantitative polymerase chain reaction (qPCR).

The *P. falciparum* 3D7 clone, produced at Queensland Institute of Medical Research in Brisbane, Australia, was used in VAC069E. The 3D7 blood stabilate was isolated from a single O-donor who was infected by mosquito bite^22^. On the day of CHMI, the cryopreserved inoculum was thawed and diluted to a target dose of 1000 parasitised erythrocytes in 5 mL normal saline for each participant and administered intravenously as previously described^21^.

Following the day of CHMI, participants were reviewed in clinic once to twice daily for symptoms of malaria and quantification of parasitaemia. Clinic visits commenced from day 1 after CHMI during VAC069A; day 6 in VAC069B; day 7 in VAC069C, VAC069D and VAC079 and day 5 after *P. falciparum* CHMI in VAC069E.

Participants received antimalarial treatment (artemether/lumefantrine or atovaquone/proguanil) if they had significant malaria symptoms and parasitaemia ≥5,000 gc/mL as measured by qPCR, or if parasitaemia reached ≥10,000 gc/mL irrespective of symptoms. Positive thick film microscopy (defined as ≥2 malaria parasites seen in 200 fields) was also included in the malaria diagnostic criteria during CHMI in VAC069A and B. In subsequent CHMIs in VAC069C-E and VAC079, microscopy was removed as a diagnostic tool due to low sensitivity. The diagnostic algorithms are detailed in the Supplementary Appendix. Participants were reviewed daily until completion of antimalarial treatment on day 3 and again at 6 days post-treatment. Further clinic visits took place on days 28, 45 and 90 after the day of CHMI during VAC069A; and on days 56 and 96 during VAC069B-E.

### Safety analysis

At each clinic visit following CHMI until day 9 after treatment, participants were asked to report any unsolicited AEs and the presence and severity of solicited AEs, consisting of a list of thirteen symptoms commonly associated with malaria infection. Unsolicited AEs were assessed for causality in relation to CHMI or antimalarial treatment by the Investigators. Antimalarial treatment associated AEs were also solicited from day 1 after initiation of treatment until day 6 after treatment. Participants graded the severity of their symptoms from 1 (mild) to 3 (severe), as per severity grading criteria shown in the Supplementary Appendix. SAEs were collected for the duration of the study. Physical observations were taken at each clinic visit and participants were asked to record their oral temperature if they experienced feverishness outside of clinic visits.

Blood samples taken 1 to 2 days before the day of CHMI, weekly during CHMI, on the day of malaria diagnosis and at post-treatment visits were tested for full blood count and biochemistry at Oxford University Hospitals NHS Foundation Trust. Biochemistry tests consisted of plasma electrolytes, urea, creatinine, bilirubin, ALT, ALP and albumin. Blood was tested for hepatitis B, hepatitis C, HIV, EBV and CMV prior to and at 3 months after each CHMI and for red blood cell alloantibodies at 3 months after each CHMI. Extensive safety testing of the PvW1 and 3D7 stabilates, including for presence of blood-borne viruses, have previously been described^19,23^.

### qPCR and modelling of parasite multiplication rate

*P. vivax* and *P. falciparum* parasitaemia was measured by qPCR in blood in real-time using an assay that targets the 18S ribosomal RNA gene, as detailed in the Supplementary Appendix^19^. The mean of three replicate qPCR results for each individual at each timepoint was used to model the PMR for each participant. PMR was calculated from the slope of a linear model fitted to log_10_ transformed qPCR data^24^.

### Processing whole blood for plasma

To obtain plasma, venous blood was drawn into K_2_EDTA-coated vacutainers (BD). 3 ml whole blood was divided into two 2 ml Eppendorf tubes and centrifuged at 1000 xg for 10 minutes at 4°C to pellet cellular components. Plasma was then carefully transferred to a new 2 ml tube and centrifuged at 2000 xg for 15 minutes at 4°C to pellet platelets. Cell-free, platelet-depleted plasma was aliquoted into 1.5 ml Eppendorf tubes, snap frozen on dry ice and stored at -80°C.

### Serum preparation

For serum preparation, blood samples were collected into untreated vacutainers and incubated at room temperature. The clotted blood was then centrifuged for 5 min at 750 x g. Serum was stored at -80°C.

### Antibody response to P. vivax antigens by ELISA and multiplex assay

Total anti-PvDBPII IgG serum concentrations were assessed over time by ELISA using standardised methodology^25^. For the multiplex assay, recombinant proteins covering the entire extracellular domains of seven *P. vivax* merozoite antigens, plus CD4 control, were expressed in mammalian cells^26^: PvAMA1, PvDBP, PvEBP, PvGAMA, PvMSP1, PvP12 and PvTRAg25 (**Table S5**). Mag-Plex Luminex beads were coupled with each antigen and a master mix containing 25 beads of each antigen per μl was prepared. Heat inactivated serum samples were diluted 1:200 and incubated with the bead master mix before addition of anti-human IgG secondary antibody. Fluorescence Intensity was measured using BioPlex 200 Systems (Bio-Rad).

### Multiplexed plasma analyte analysis

The concentration of 24 analytes was measured in plasma samples collected at baseline, during malaria infection, at diagnosis, 1, 3 and 6 days after drug treatment and at 45 to 96 days post-CHMI. For this analysis we selected 10 participants who had completed at least 2 CHMIs: 7 participants who underwent primary *P. vivax* CHMI in VAC069C and secondary *P. vivax* CHMI in VAC069D, as well as 3 participants who underwent primary *P. vivax* CHMI in VAC069D. Of these 10 participants, 6 underwent heterologous rechallenge with *P. falciparum* during VAC069E. Plasma was thawed on ice and centrifuged at 1000 xg for 1 minute at 4°C to remove potential protein aggregates. We customised 2 LEGENDplex panels from BioLegend and performed each assay on filter plates according to the manufacturer s instructions. We included the acute phase proteins C reactive protein (CRP), pentraxin 3 (PTX3) and serum amyloid A (SAA); pyrogenic cytokines and their regulators IL-1RA, IL-6 and sTNFRII; interferon gamma (IFNγ), IL-10 and the endogenous alarmin calprotectin (MRP8/14) for immune regulation; myeloperoxidase (MPO) - an enzyme secreted by activated neutrophils and associated with extracellular traps; CXCL10)which promotes T cell recruitment into inflamed tissues; IL-12p70 which polarises CD4 T cells towards an inflammatory T_H_1 fate; IL-15 and IL-18 which can induce bystander T cell activation; markers of cytotoxicity including granzyme B (GZMB) and perforin; myoglobin to indicate collateral tissue damage; matrix metalloproteases (MMP) 9 and 10 which can degrade extracellular matrix to aid tissue remodelling; E-selectin (endothelial cell adhesion molecule), ICAM-1 and VCAM -1 (intercellular / vascular cell adhesion molecules) as markers of endothelium activation and dysfunction; and markers associated with coagulation (D-Dimer and tissue plasminogen activator (tPA)). Samples and standards were acquired on an LSRFortessa flow cytometer (BD) and FCS files were processed using LEGENDplex software (version 2023-02-15 (49495)), which automatically interpolates a standard curve using the plate-specific standards and calculates analyte concentrations for each sample. Downstream data analysis was performed in R using the ggplot2^27^ and ComplexHeatmap^28^ packages for plotting.

### Statistical analysis

Data were analysed using R (v4.4.2). To determine if haematological / biochemical parameters and plasma proteins varied significantly between primary and secondary *P. vivax* infection at each time-point, we performed linear regression using the lme4 package to fit mixed-effects models that included time-point and infection number as categorical fixed effects and participant as a random effect. For heterologous *P. falciparum* CHMI we tested secondary *vivax* versus *falciparum*. In every case, linear hypothesis testing was performed using multcomp’s glht function with Benjamini-Hochberg correction for multiple testing across all analytes. Pairwise comparisons between two groups at a single time-point were performed using Wilcoxon signed-rank test utilising wilcox.test from the stats package in R or GraphPad Prism version 9.5.0 (GraphPad Software Inc). A value of p <0.05 was considered significant.

## Data Availability

All data produced in the present study are available upon reasonable request to the authors

## Acknowledgments

The authors are grateful for the assistance of Aabidah Ali, Duncan Bellamy, Nicholas Byard, Federica Cappuccini, Hannah Davies, Amy Flaxman, Julie Furze, Michelle Fuskova, Daniel Jenkins, Kimberly Johnson, Kathryn Jones, Reshma Kailath, Colin Larkworthy, Alison Lawrie, Meera Madhavan, Rebecca Makinson, Daniel Marshall-Searson, Ruth Payne, Jack Quaddy, Indra Rudiansyah, Hannah Scott, Iona Taylor, Cheryl Turner, Nicola Turner, Marta Ulaszewska, Chris Williams, Rhea Zambellas (Jenner Institute, University of Oxford); Sally Pelling-Deeves for arranging contracts (University of Oxford); Julie Staves and the Haematology Laboratory (Oxford University Hospitals NHS Foundation Trust); Nongnuj Maneechai, Tianrat Piteekan, Nattawan Rachaphaew, Wanlapa Roobsoong, Jetsumon Sattabongkot, Nick Day (Mahidol Vivax Research Unit, Thailand); members of the MultiViVax Scientific Advisory Board; and all of the participants who participated in these trials.

---

**Figure S1.**
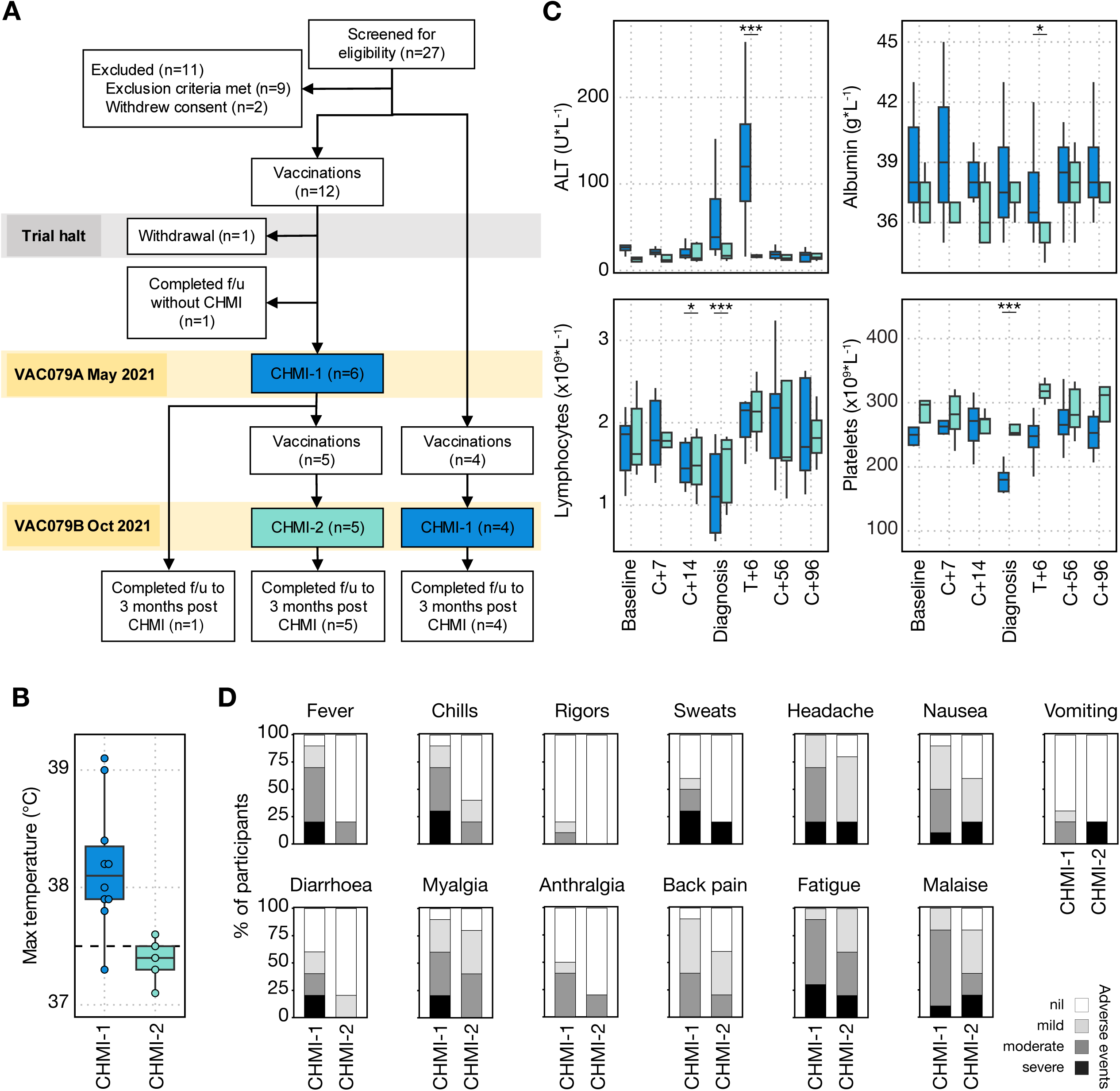
Primary and secondary *P. vivax* CHMI in the VAC079 study. **(A)** VAC079 tested the efficacy of a *P. vivax* PvDBPII protein/adjuvant vaccine. 10 participants completed 3 vaccinations prior to undergoing primary *P. vivax* CHMI (CHMI-1) at 2 to 4 weeks after their third vaccination. Five participants received a fourth vaccination, followed by secondary homologous *P. vivax* CHMI (CHMI-2). **(B)** Maximum recorded core temperature during primary and secondary CHMI (*p* = 0.1 by Wilcoxon signed-rank test). **(C)** Alanine aminotransferase (ALT), serum albumin, lymphocyte and platelet counts were measured in peripheral blood during primary and secondary CHMI at baseline (1 or 2 days before challenge); 7 and 14 days after challenge (C+7 and C+14); on the day of diagnosis; 6 days after treatment (T+6); and 56 (C+56) and 96 days (C+96) after challenge. Statistically significant differences between CHMI-1 and CHMI-2 were identified at each time-point by using mixed-effects modelling and linear regression (* *p* < 0.05 and *** *p* < 0.001). Box and whisker plots show median and IQR (whiskers represent 1.5x IQR). **(D)** Clinical signs and symptoms of malaria in participants undergoing primary and secondary CHMI in the VAC079 study. Data show the maximum severity of each solicited adverse event (AE), consisting of 13 malaria symptoms, reported by an individual in the 48 hours before and after diagnosis as a proportion of the total number of participants undergoing CHMI. In (B-D) n = 10 (primary *P. vivax* CHMI) and n = 5 (secondary *P. vivax* CHMI).

**Figure S2.**
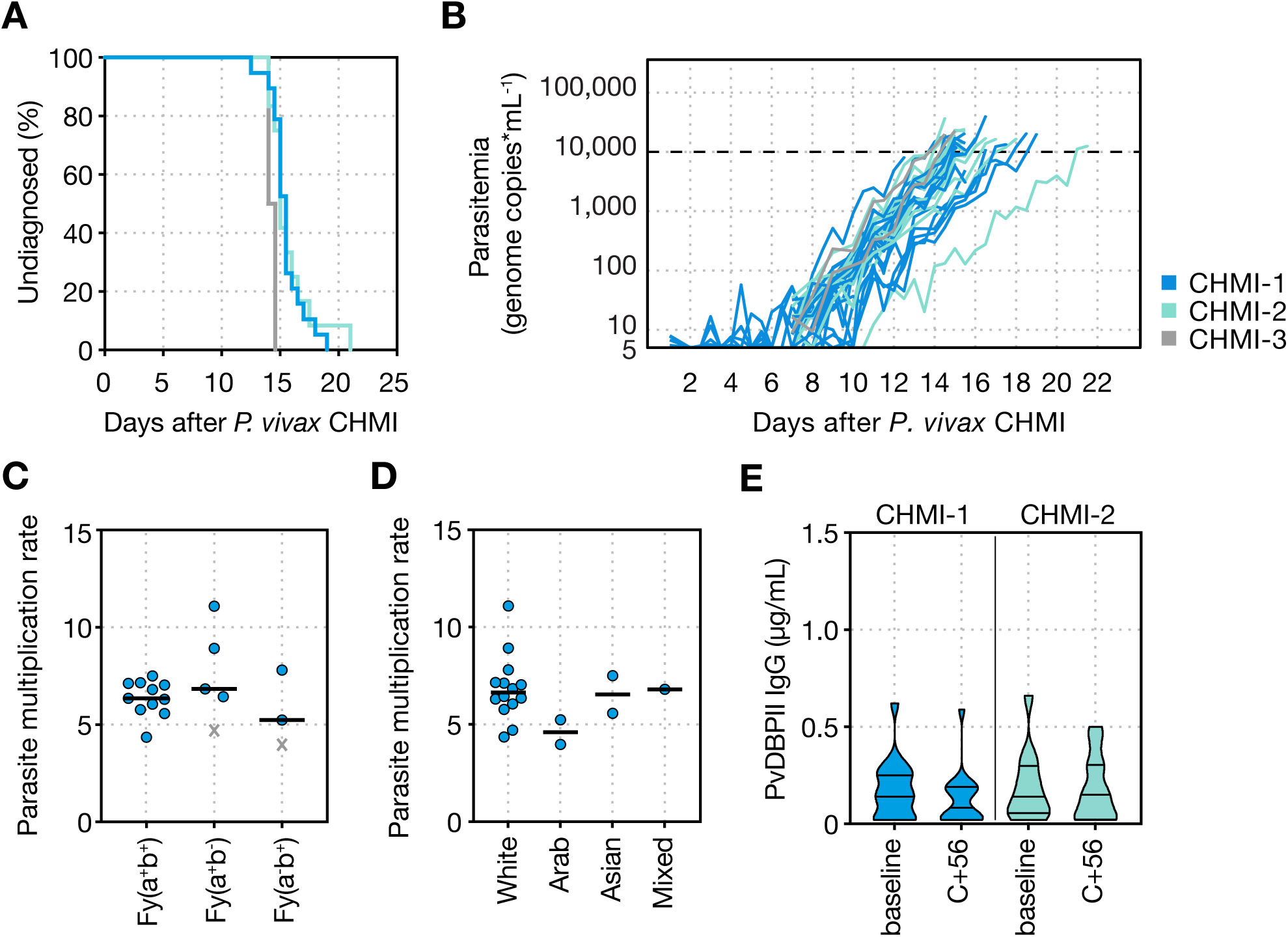
Parasite growth during *P. vivax* CHMI in the VAC069 study. **(A)** Kaplan-Meier plot showing the time to malaria diagnosis in the VAC069 study. Median time to diagnosis was 15.5 days during primary (CHMI-1) and 15.0 days during secondary *P. vivax* CHMI (CHMI-2); no significant difference by log-rank test (p = 0.8). Median time to diagnosis was 14.25 days during tertiary CHMI (CHMI-3) **(B)** Parasitaemia was measured up to twice daily by qPCR, note the two participants during tertiary *P. vivax* (CHMI-3, shown in grey). The dashed line indicates the treatment threshold of 10,000 gc ml^-1^. **(C)** Parasite multiplication rate (fold-change per 48 hours) during primary CHMI is grouped by Duffy phenotype; no significant difference between group medians by Kruskal-Wallis test. The two individuals designated by a cross were heterozygous for the erythroid silent FY*B allele on Duffy genotyping. **(D)** Parasite multiplication rate (fold-change per 48 hours) during primary CHMI is grouped by participant ethnicity. **(E)** Violin plot shows anti-PvDBPII IgG serum concentrations as measured by ELISA at 1 or 2 days before challenge (baseline) and 56 days after challenge (C+56) during primary (CHMI-1) and secondary *P. vivax* CHMI (CHMI-2). All responses are below the limit of quantification (1 μg ml^-1^). In (A-D) n = 19 (primary *P. vivax* CHMI) and n = 12 (secondary *P. vivax* CHMI).

**Figure S3.**
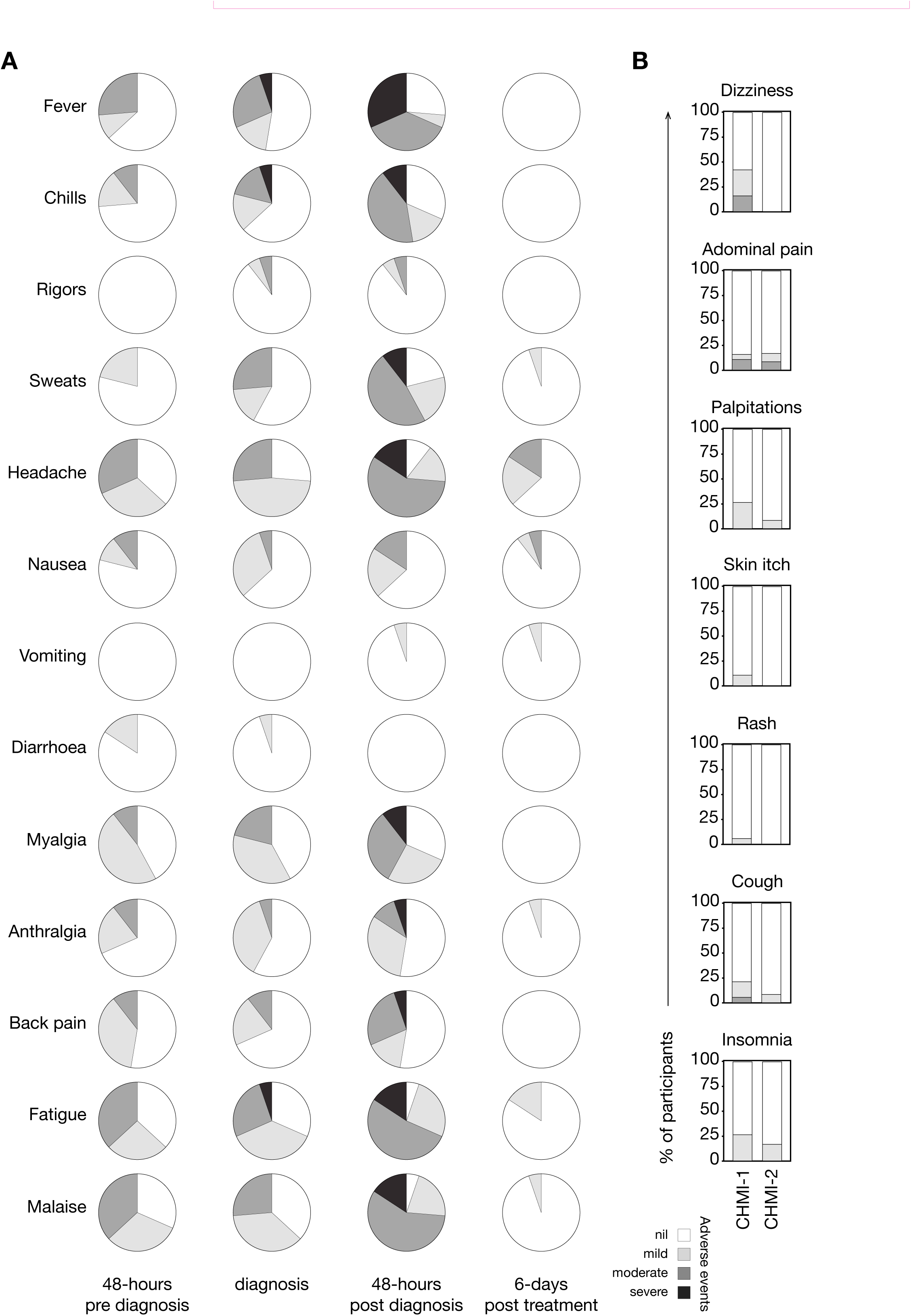
Solicited adverse events during *P. vivax* CHMI in the VAC069 study. **(A)** Clinical signs and symptoms of malaria in participants undergoing primary *P. vivax* CHMI in the VAC069 study. Data show the maximum severity of each solicited adverse event (AE), consisting of 13 malaria symptoms, as reported by an individual in the 48 hours before diagnosis; at diagnosis; the 48 hours after diagnosis; and 6 days after starting antimalarials. All data are shown as a proportion of the total number of participants undergoing CHMI (n = 19). **(B)** Solicited adverse events (AE) associated with antimalarial treatment during primary (CHMI-1, n = 19) and secondary (CHMI-2, n = 12) *P. vivax* CHMI. The maximum severity of each AE is shown as a proportion of the total number of participants undergoing CHMI.

**Figure S4.**
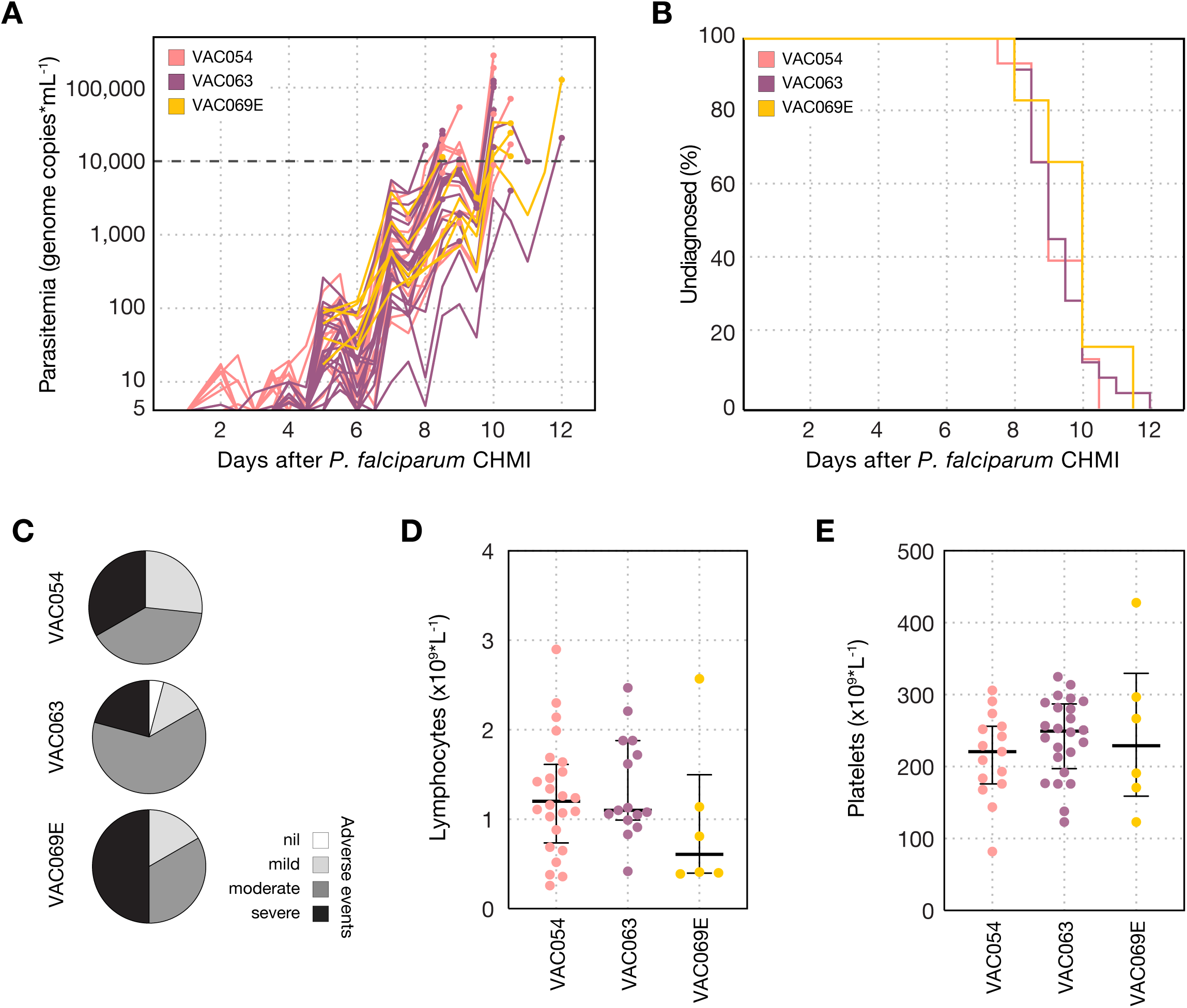
Parasitaemia and solicited adverse events during heterologous rechallenge are similar to primary *P. falciparum* CHMI. **(A)** Parasitaemia was measured up to twice daily by qPCR during heterologous rechallenge with *P. falciparum* (VAC069E) and compared to primary CHMI in malaria-naïve participants in two of our previous CHMI studies (VAC054 and VAC063), in which participants were inoculated with red bloods cells infected with the same 3D7 clone of *P. falciparum* ^21,33,34^. The diagnostic criteria used in VAC054 and the first cohort of VAC063 (VAC063A) combined microscopy and qPCR, whereas in VAC063B-C and VAC069E only qPCR was used. Despite this no significant differences in the parasitaemia at diagnosis were seen across all CHMIs. The dashed line indicates the treatment threshold for VAC069E. **(B)** Kaplan-Meier plot showing the time to malaria diagnosis in VAC069E compared to primary CHMI in VAC054 and VAC063. Median time to diagnosis was 10 days in VAC069E and 9 days in both VAC054 and VAC063; no significant difference by log-rank test (*p* = 0.6). **(C)** Maximum severity of any solicited adverse event (AE), consisting of 13 malaria symptoms, reported by an individual in the 48 hours before and after diagnosis during primary *P. falciparum* CHMI in VAC054, VAC063 compared to VAC069E. Data are shown as a proportion of the total number of participants undergoing each CHMI. **(D)** Lymphocyte and **(E)** platelet counts were measured in peripheral blood on the day of diagnosis in VAC069E and compared to primary *P. falciparum* CHMI in VAC054 and VAC063. No significant differences between groups by Kruskal-Wallis test (lymphocytes *p* = 0.3, platelets *p* = 0.5). Median and IQR shown. In (A-E) n = 15 (VAC054), n = 24 (VAC063), n = 6 (VAC069E).

