## Supplemental methods and tables for "An experimental model of clinical immunity for human malaria"

### Supplementary Methods

#### VAC069 Study Inclusion and Exclusion Criteria

A medical history and physical examination were conducted at the screening visit, as well as baseline blood tests including a full blood count; urea and electrolytes; liver function tests; and hepatitis B virus (HBV), hepatitis C virus (HCV), human immunodeficiency virus (HIV), Epstein-Barr virus (EBV) and Cytomegalovirus (CMV) serology. Dipstick urinalysis for all volunteers and urinary pregnancy testing for all female volunteers were also conducted at screening. Pregnancy testing (serum Beta Human Chorionic Gonadotropin, Beta hCG), was also carried out prior to and during CHMI. A full list of inclusion and exclusion criteria is shown below for the VAC069 study:

The volunteer must satisfy all the following criteria to be eligible for the study:

- Healthy adult aged 18 to 50 years.
- Red blood cells positive for the Duffy antigen/chemokine receptor (DARC).
- Normal serum levels of Glucose-6-phosphate dehydrogenase (G6PDH).
- Negative haemoglobinopathy screen.
- Able and willing (in the Investigator’s opinion) to comply with all study requirements.
- Willing to allow the Investigators to discuss the volunteer’s medical history with their General Practitioner.
- Volunteers with the potential to become pregnant only: Must practice continuous effective contraception* for the duration of the clinic visits (first 3 months post-CHMI).
- Agreement to permanently refrain from blood donation.
- Written informed consent to participate in the trial.
- Reachable (24/7) by mobile phone during the period between CHMI and completion of all antimalarial treatment.
- Willing to take a curative anti-malarial regimen following CHMI.
- Willing to reside in Oxford for the duration of the study, until antimalarials have been completed.
- Answer all questions on the informed consent quiz correctly.

* Female volunteers are required to use an effective form of contraception during the course of the study as malaria challenge could pose a serious risk to both maternal health and the unborn foetus.

Acceptable forms of contraception for female volunteers include:

- Established use of oral, injected or implanted hormonal methods of contraception (an additional form of contraception will be required when taking the antimalarial medication, as this can interfere with the efficacy of hormonal contraception).
- Placement of an intrauterine device (IUD) or intrauterine system (IUS).
- Total abdominal hysterectomy.
- Barrier methods of contraception (condom or occlusive cap with spermicide).
- Male sterilisation, if the vasectomised partner is the sole partner for the subject.
- True abstinence, when this is in line with the preferred and usual lifestyle of the subject (periodic abstinence and withdrawal are not acceptable methods of contraception).

The volunteer may not enter the study if any of the following apply:

- History of clinical malaria (any species).
- Travel to a clearly malaria endemic locality during the study period or within the preceding six months.
- Use of systemic antibiotics with known antimalarial activity within 30 days of CHMI (e.g. trimethoprim-sulfamethoxazole, doxycycline, tetracycline, clindamycin, erythromycin, fluoroquinolones and azithromycin).
- Haemoglobin <120 g/L for a female volunteer or <130 g/L for a male volunteer prior to primary CHMI. (However, for enrolment into secondary and tertiary CHMIs slightly lower haemoglobin values (≤0.5 g/L) will be permitted at the discretion of the Investigator, to account for the blood volume donated during the previous CHMI).
- Receipt of immunoglobulins within the three months prior to enrolment.
- Receipt of blood transfusion at any time in the past.
- Peripheral venous access unlikely to allow twice daily blood testing (as determined by the Investigator).
- Receipt of an investigational product in the 30 days preceding enrolment, or planned receipt during the study period.
- Prior receipt of an investigational vaccine likely to impact on interpretation of the trial data or the *P. vivax* parasite as assessed by the Investigator.
- Planned receipt of a COVID-19 vaccine between 2 weeks before the day of CHMI until completion of antimalarial treatment
- Any confirmed or suspected immunosuppressive or immunodeficient state, including HIV infection; asplenia; recurrent, severe infections and chronic (more than 14 days) immunosuppressant medication within the past 6 months (inhaled and topical steroids are allowed).
- History of allergic disease or reactions likely to be exacerbated by malaria infection.
- Pregnancy, lactation or intention to become pregnant during the study.
- Use of medications known to cause prolongation of the QT interval ***and*** existing contraindication to the use of Malarone.
- Use of medications known to have a potentially clinically significant interaction with Riamet ***and*** Malarone.
- Any clinical condition known to prolong the QT interval.
- History of cardiac arrhythmia, including clinically relevant bradycardia.
- Disturbances of electrolyte balance, e.g. hypokalaemia or hypomagnesaemia.
- Family history of congenital QT prolongation or sudden death.
- Contraindications to the use of both of the proposed anti-malarial medications Riamet and Malarone.
- History of cancer (except basal cell carcinoma of the skin and cervical carcinoma in situ).
- History of serious psychiatric condition that may affect participation in the study.
- Any other serious chronic illness requiring hospital specialist supervision.
- Suspected or known current alcohol abuse as defined by an alcohol intake of greater than 25 standard UK units every week.
- Suspected or known injecting drug abuse in the 5 years preceding enrolment.
- Hepatitis B surface antigen (HBsAg) detected in serum.
- Seropositive for hepatitis C virus (antibodies to HCV) at screening or at C-7 (***unless*** has taken part in a prior hepatitis C vaccine study with confirmed negative HCV antibodies prior to participation in that study, and negative HCV RNA PCR at screening for this study).
- Positive family history in both 1^st^ AND 2^nd^ degree relatives < 50 years old for cardiac disease.
- Volunteers unable to be closely followed for social, geographic or psychological reasons.
- Any clinically significant abnormal finding on biochemistry or haematology blood tests, urinalysis or clinical examination. In the event of abnormal test results, confirmatory repeat tests will be requested. Procedures for identifying laboratory values meeting exclusion criteria are shown in Appendix A.
- Any other significant disease, disorder, or finding which may significantly increase the risk to the volunteer because of participation in the study, affect the ability of the volunteer to participate in the study or impair interpretation of the study data.
- Inability of the study team to contact the volunteer’s GP to confirm medical history and safety to participate.

Additional exclusion criteria for participants in VAC069C-E:

- Body weight <50 kg, as measured at screening

#### Severity grading of adverse events (AE)

Participant reported AEs were graded as mild, moderate or severe according to the following criteria:

- **GRADE 0:** None.
- **GRADE 1:** Transient or mild discomfort (< 48 h); no medical intervention/therapy required.
- **GRADE 2:** Mild to moderate limitation in activity – some assistance may be needed; no or minimal medical intervention/therapy required.
- **GRADE 3:** Marked limitation in activity, some assistance usually required; medical intervention/therapy required; hospitalization possible.

#### Causality Assessment

For each unsolicited AE, an assessment of the relationship of the AE to the study intervention(s) was undertaken. Alternative causes of the AE, such as the natural history of pre-existing medical conditions, concomitant therapy, other risk factors and the temporal relationship of the event to CHMI/ antimalarials were considered. The likely causality of all unsolicited AEs was assessed as per the criteria below:

- **No Relationship:** No temporal relationship to study intervention ***and*** alternate aetiology (clinical state, environmental or other interventions); ***and*** does not follow known pattern of response to study intervention.
- **Unlikely:** Unlikely temporal relationship to study intervention ***and*** alternate aetiology likely (clinical state, environmental or other interventions) ***and*** does not follow known typical or plausible pattern of response to study intervention.
- **Possible:** Reasonable temporal relationship to study intervention; ***or*** event not readily produced by clinical state, environmental or other interventions; ***or*** similar pattern of response to that seen with other similar interventions.
- **Probable:** Reasonable temporal relationship to study intervention; ***and*** event not readily produced by clinical state, environment, or other interventions ***or*** known pattern of response seen with other similar interventions.
- **Definite:** Reasonable temporal relationship to study intervention; ***and*** event not readily produced by clinical state, environment, or other interventions; ***and*** known pattern of response seen with other similar interventions.

#### **Severity grading criteria for clinically significant laboratory abnormalities**

Adapted from FDA guidelines using Oxford University Hospitals NHS Foundation Trust laboratory reference ranges.

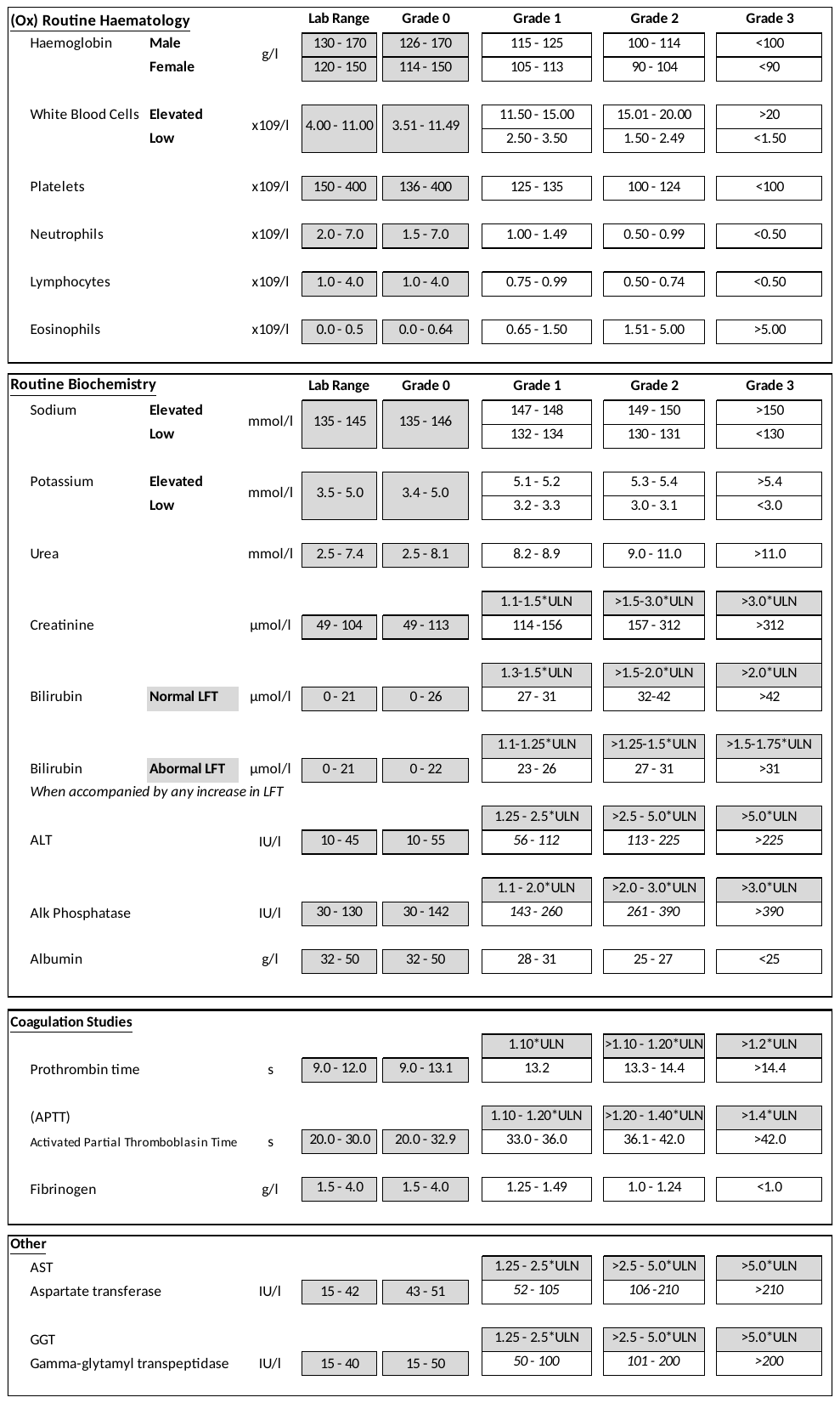

#### Multiplex immunoassay

**Recombinant protein expression and purification**

Recombinant proteins covering the entire extracellular domain of seven *Plasmodium vivax* antigens and CD4 used as a control were expressed (details of constructs in **Table S5**). The PvEBP (KMZ83376.1) construct was generously provided by Prof. Gavin Wright, while Pv12 (PVX_113775), PvMSP1 (PVX_099980), PvGAMA (PVX_088910), and PvDBP (PVX_110810) were previously utilised in a study by Hostetler et al. ^1^, with plasmids sourced from Addgene and subcloned into the pTT3 expression plasmid, containing a Biolinker peptide and a C-terminal His tag. The extracellular domains of PvAMA1 (PVP01_0934200) and PvTRAg25 (PVP01_0000100) were determined using signal peptide analysis ^2^ and transmembrane prediction servers ^3^. The designed constructs, with codons optimized based on the human genome and all predicted N-glycosylation sites (NXS/T) were mutated by converting the S/T to A in order to prevent inappropriate glycosylation events (*Plasmodium* parasites do not synthesise N-linked glycans). Constructs were synthesised by TWIST Biosciences, UK and cloned into a mammalian expression plasmid with a 5’ mouse variable κ light chain signal peptide and an enzymatic biotinylation sequence/hexa-His tag at the C-terminus. PvDBP (PVX_110810) additionally incorporates a CD4 tag at the C-terminus. Plasmids were amplified, purified, and diluted to 1 mg/ml, then mixed with polyethyleneimine (PEI 40 kDa) in a 1:2.5 ratio for transfection in a 50 ml culture. The DNA/PEI mixture was incubated for 8 min before being added to HEK 293E cells cultured in Freestyle 293 media supplemented with Geneticin and 1% fetal bovine serum. Cell maintenance involved shaking at 125 rpm, 37°C, 5% CO_2_, and 70% relative humidity.

To purify proteins from the culture supernatant, HEK 293E cells were pelleted at 3000 g for 20 min and subsequently filtered (0.22 µm, Corning). The culture supernatant was adjusted to 5 mM imidazole and 300 mM NaCl before being loaded onto Talon Sepharose, a cobalt-based immobilised metal affinity chromatography (IMAC) resin. The Talon Sepharose resin was initially equilibrated with binding buffer (50 mM sodium phosphate, 300 mM NaCl, pH 7.4), before culture supernatant was passed through the column to allow for protein binding to the beads. The column was washed with 20 ml of wash buffer (50 mM sodium phosphate, 300 mM NaCl, 5 mM imidazole, pH 7.4), before recombinant proteins were eluted with 5 ml of elution buffer (50 mM sodium phosphate, 300 mM NaCl, 150 mM imidazole, pH 7.4). The eluted fraction was subjected to buffer exchange with PBS using 10 kDa Vivaspin Ultracel concentrators. Final concentrations were determined using a Nanodrop OneC UV–Vis Spectrophotometer (Thermo Fisher), and the purified proteins were run in 12% SDS PAGE to assess purity and stored at 4 °C until coupling.

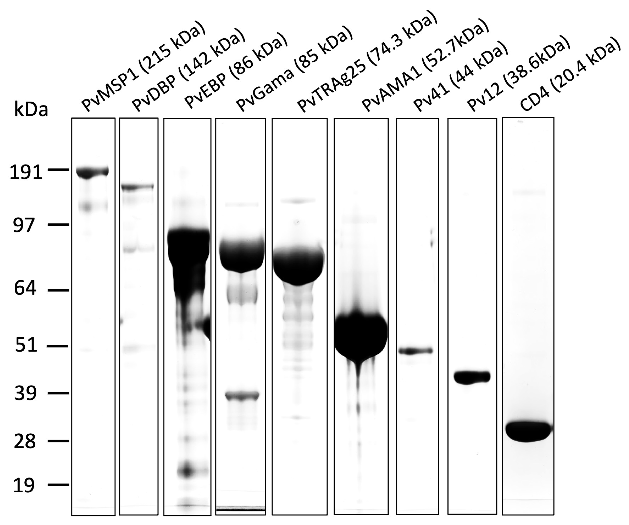

**Protein production for multiplex immunoassay.** Affinity-purified proteins were analysed by 12% SDS-PAGE to assess purity, with corresponding molecular weights indicated.

**Protein conjugation to the Mag-Plex Luminex beads**

Bead stocks (2.5x10^6^/ml, Luminex, MC100XX-YY) were dispersed using a bath sonicator (30 s pulse, 10 s off). Subsequently, 0.4 ml of the bead stock (1x10^6^/ml) was dispensed into low bind Eppendorf tubes, pelleted, and the supernatant was removed using a bar magnet. The beads underwent a wash with 1 ml sterile water (Sigma, W3500-500ml). A stock solution of EDC (1-ethyl-3(-3-dimethylaminopropyl) carbodiimide hydrochloride) and NHS (N-hydroxysuccinimide) was prepared in activation buffer (0.1 M NaH_2_PO_4_, pH 6.2) and 50 μl of EDC (Thermo, 22980) (50 mg/ml) and NHS (Thermo, 24500) (50 mg/ml) were added to the beads and vortexed gently. After a 20 min incubation at 25 °C on a rotator at 40 rpm, the supernatant was removed, and the beads were washed twice with 50 mM MES (4-morpholinoethanesulfonic acid) pH 5.0. Ten micrograms of each protein were added to the beads, and the reaction volume was adjusted to 1 ml with MES pH 5.0. The reaction mixture was incubated for 2 hours at 25 °C on a rotator at 40 rpm. Following incubation, the bead-coupled proteins underwent three washes with PBS-TBN (PBS 1X, 0.1% BSA, 0.05% Tween20, 0.05% sodium azide) and were finally resuspended in 1ml of PBS-TBN. Bead concentration was measured in a cell counter, and the beads were stored at 4 °C in the dark until further use. All steps were conducted in the dark, and magnetic base washing facilitated the separation of the beads.

The coupling of the beads was validated using flow cytometry. Approximately 10,000 antigen-coupled beads were aliquoted into an Eppendorf tube, and 5 μl of PE-conjugated Anti-his Ab (R&D system, IC050P) was added to each tube. The reaction mixture was incubated for 2 hours at 4 °C, followed by centrifugation at 12,000 rpm for 1 min and washing with PBS-TBN (this step was repeated twice). The beads were finally resuspended in PBS-TBN and analysed by flow cytometry (Thermo, Attune) with YL1 laser (561nm) for PE (561 585/16, laser power 400).

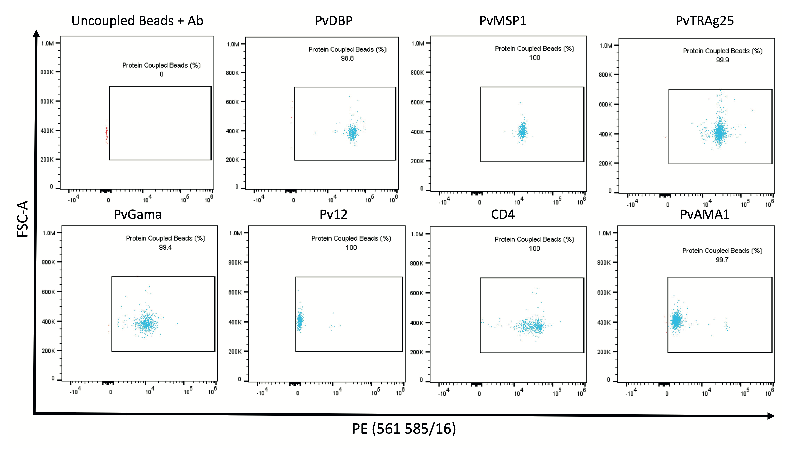

**Protein bead conjugation for multiplex immunoassay.** Flow cytometry dot plots validating protein bead conjugation. The Y-axis represents forward scatter area, while the X-axis displays the fluorescence signal from the anti-His antibody PE conjugate. Protein-coupled beads are depicted in the gated population (sky blue) in contrast to the uncoupled beads (red).

**Luminex Assay**

Each bead set underwent sonication and vortexing, and a master mix containing 25 beads of each region/μl was prepared in PBS-TBN. The total volume of the Bead Master mix, amounting to 12 ml, was generated by adding the required quantity of each protein-coupled bead for eight antigens, and the remaining volume was topped up with PBS-TBN. After another round of sonication and vortexing, 50 μl of the master mix was dispensed into each well in duplicate. The bead calculations are detailed in **Table S5**.

Human serum samples were heat-inactivated at 56 °C for 30 minutes, and a master plate was prepared with sera diluted at 1:200 in PBS-TBN. Subsequently, 50 μl of serum samples was added to plates containing 50 μl of beads, mixed five times, covered with aluminium foil, and incubated on a plate shaker at 200 rpm for 1 h at 25 °C. Following incubation, the plate was fixed on a magnetic base and washed five times (5 x 100 μl/well) with PBS-TBN, with each wash lasting 1 min. Fifty microliters of goat anti-human IgG secondary antibody (4 μg/ml) (Thermo, PA1-86078 ) was added to each well and incubated for 30 min at 25 °C. The plates were washed five times (5 x 100 μl) with PBS-TBN, fixed on a magnetic base, and finally resuspended in 100 μl PBS-TBN. Prior to the run, calibration of the instrument (Bio-Rad, BioPlex 200 Systems) was performed, and the plates were shaken for 5 min. A blank sample well, containing only the bead master mix, was included in the plate to subtract any background fluorescence from the data.

#### Anti-PvDBPII standardized enzyme-linked immunosorbent assay (ELISA)

ELISAs to quantify circulating PvDBPII-specific total IgG responses were performed using standardised methodology, similar to that previously described ^4^. Nunc MaxiSorp ELISA plates (Thermo Fisher) were coated overnight (≥16 h) at 4 °C with 50 µL per well of 2 µg/mL PvDBPII protein. Plates were washed 6x with 0.05 % phosphate-buffered saline/Tween (PBS/T) and tapped dry. Plates were blocked for 1 h with 100 µL per well of Starting Block T20 (Thermo Fisher) at 20 °C. Test samples were diluted in blocking buffer (minimum dilution of 1:100), and 50 µL per well was added to the plate in triplicate. Reference serum (made from a pool of high-titer vaccinated donor serum) was diluted in blocking buffer in a three-fold dilution series to form a ten-point standard curve. Three independent dilutions of the reference serum were made to serve as internal controls. The standard curve and internal controls were added to the plate at 50 µL per well in duplicate. Plates were incubated for 2 h at 20 °C and then washed 6x with PBS/T and tapped dry. Goat anti-human IgG–alkaline phosphatase secondary antibody (Merck) was diluted 1:1000 in blocking buffer and 50 µL per well was added. Plates were incubated for 1 h at 20 °C. Plates were washed 6x with PBS/T and tapped dry. 100 µL per well of p-nitrophenyl phosphate alkaline phosphatase substrate (Thermo Fisher) was added, and plates were incubated for approximately 15 min at 20 °C. Optical density at 405 nm (OD_405_) was measured using an ELx808 absorbance reader (BioTek) until the internal control reached an OD_405_ of 1.0. The reciprocal of the internal control dilution giving an OD_405_ of 1.0 was used to assign an arbitrary unit (AU) value of the standard. Gen5 ELISA software v3.04 (BioTek) was used to convert the OD_405_ of test samples into AU by interpolating from the linear range of the standard curve fitted to a four-parameter logistic model. Any test samples with an OD_405_ below the linear range of the standard curve at the minimum dilution tested were assigned a minimum AU value of 5.0. These responses in AU are reported in μg/mL of PvDBPII following generation of a conversion factor by calibration-free concentration analysis (CFCA). In short, CFCA was performed using a Biacore X100 instrument, a Biotin CAP chip and X100 control and evaluation software (Cytiva). Purified mono-biotinylated antigen was produced for use in CFCA and chip was regenerated with manufacturer’s supplied regeneration and CAP reagents and fresh antigen prior to each application of antibody. Serum samples, from a previous clinical trial (VAC051), with a range of PvDBP antibody responses were diluted and assessed for antigen-specific antibody binding and initial rates of antigen-specific binding at 5 μL/min and 100 μL/min measured and compared to permit measurement of concentration. The CFCA-measured PvDBP-specific antibody concentrations for each individual were analysed by linear regression with corresponding total IgG ELISA AU data, where slope of the line was used to derive an AU-to-μg/mL conversion factor.

#### Blood-stage inoculum preparation and CHMI

The *P. vivax* W1 clone blood-stage inoculum was thawed and prepared under strict aseptic conditions as previously described ^5^. The required number of vials of the cryopreserved stabilate (each containing approximately 0.5 mL of red blood cells in 1 mL of Glycerolyte 57) were thawed in parallel in an area using solutions licensed for clinical use and single-use disposable consumables. A class II microbiological safety cabinet (MSC) was used to prepare the inoculum, which was fumigated with hydrogen peroxide and decontamination validated prior to use. To prepare the inoculum, 0.2 volume 12 % saline was added dropwise to the contents (about 1.5 mL) of each vial of thawed infected blood. Each sample was left for 5 min, before an additional 10 volumes of 1.6 % saline was added dropwise prior to centrifugation for 4 min at 830 x *g*. Each supernatant was removed and 10 mL of 0.9 % saline was added dropwise. The cell pellets were pooled and washed twice in 0.9 % saline before a final resuspension into one 10 mL sample in 0.9 % saline. This 10 mL suspension was then divided into aliquots, equivalent to one tenth of one original cryovial. Each aliquot was made up to a total volume of 5 mL in 0.9 % saline in a sterile syringe for injection and transported to the clinic. For each challenge, one dose of the 1:10 diluted inoculum was quantified by quantitative polymerase chain reaction (qPCR) to be equivalent to between 165 to 217 genome copies of *P. vivax*. This will be an overestimate of the number of live viable parasites administered per participant because some parasites will be killed during the inoculum thawing and preparation process.

The reconstituted blood-stage inoculum (5 mL per syringe) was injected intravenously using an indwelling cannula, preceded and followed by a saline flush. The inoculum was administered to all participants within a maximum of 3 h 7 min from thawing of the inoculum. Participants were observed for 1 h following injection of the inoculum before discharge from the clinical facility. Following each CHMI, a leftover sample of the inoculum was cultured and shown to be negative for bacterial contamination.

For *P. falciparum* CHMI, cryopreserved blood-stage *P. falciparum* (3D7 clone) parasites were thawed and prepared under aseptic conditions using the similar methods as for *P. vivax* inoculum preparation and as previously described^6^. The target inoculum dose for *P. falciparum* inoculum was 1000 parasitised erythrocytes per participant, which was based on microscopic estimates of the donor’s parasite density prior to freezing of the inoculum. Volunteers received infected red cells in a total volume of 5 mL normal saline, followed by a saline flush.

#### Malaria parasite quantification by qPCR

Quantitative (q)PCR was used to measure *P. vivax* parasitemia in participants’ blood in real-time using an assay that targets the 18S ribosomal RNA (rRNA) gene as previously described^5^. DNA was extracted from 0.4 mL whole EDTA blood using a QIAsymphony SP robot, utilising the Qiagen DSP Blood Midi Kit and the pre-loaded Blood 400 v6 extraction protocol, with a 100 μL elution in ATE buffer selected. Additionally, aliquots of baseline samples taken within 2 days pre-CHMI were spiked with a known concentration of positive control DNA to check there was no presence of PCR inhibitors in participants’ blood prior to CHMI.

Following DNA extraction, a standard Taqman absolute quantitation was used against a standard curve to amplify a 183 bp PCR product from the multi-copy, highly conserved 18S ribosomal RNA genes of *Plasmodium spp.* qPCR used the following adapted oligonucleotide primers and probe^7^: 18s forward primer 5’-AGG AAG TTT AAG GCA ACA ACA GGT-3’, 18s reverse primer 5’-GCA ATA ATC TAT CCC CAT CAC GA-3’ and shortened FAM labelled probe sequence 5’-TGA ACT AGG CTG CAC GCG-3’, was run on an ABI StepOne Plus machine with v2.3 software. Default Universal qPCR (target FAM-NFQ-MGB) and quality control (QC) settings were used apart from the use of 40 cycles and 25 μL reaction volume.

This qPCR detects DNA from pan-*Plasmodium* species, but unlike the synchronous growth of *P. falciparum*, circulating *P. vivax*-infected red blood cells may contain up to 10 to 15 individual genomes (in blood-stage late trophozoites and schizonts) and can also include the presence of gametocytes. The qPCR score is therefore reported in genome copies/mL (gc/mL) as opposed to a quantity of parasites.

The standard curve was generated from dilution of a linearised plasmid encoding part of the *Plasmodium spp.* 18S ribosomal RNA gene and calibrated using known *P. falciparum* (Pf) spiked blood samples initially and then reference DNA extracted from whole blood from *P. vivax*-infected patient samples in Thailand where parasites had been quantified by microscopy (kindly provided by Mahidol University). Based upon earlier results obtained using dilution series of microscopically-counted cultured Pf parasites, a Pf-specific 18S rRNA Taqman qPCR showed a lower limit of quantification (LLQ, defined as % covariance [CV] <20%) of around 20 Pf parasites (p)/mL blood^8^. Counted parasite dilution series results also suggested that the lower limit of probable detection (LLD, that is a probability of >50% of ≥1 positive result among three replicate qPCR reactions) is in the region of 5 p/mL, whereas samples at 1 p/mL are consistently negative (24/24 qPCR reactions). Positive results in this assay (even at very low detection) are thus essentially 100 % specific for genuine parasitemia, with positive results beneath the LLQ likely to signify parasitemia in the range of 2 to 20 p/mL. Similar sensitivity in terms of genome copy detection was observed when using the pan-*Plasmodium* qPCR described above and the diluted *P. vivax*-infected patient blood test samples from Thailand. As noted, these samples had microscopically mixed life stages with varying copies of the 18S rRNA gene and thus the assay readout is reported in terms of gc/mL. Based on this and the above experiments, 20 gc/mL was set as the lower limit of detection to meet positive reporting criteria, but all raw data are shown in the Results.

For quantification of *P. falciparum* parasitemia in VAC069E, a similar qPCR method was used. DNA was extracted from 0.4 mL whole EDTA blood using the QIAamp DSP DNA Blood Mini Kit (Qiagen), with a 100 μL elution in AE buffer. Additionally, aliquots of baseline samples taken within 2 days pre-CHMI were spiked with a known concentration of positive control DNA to check there was no presence of PCR inhibitors in participants’ blood prior to CHMI.

Following DNA extraction, a standard Taqman absolute quantitation was used against a standard curve to amplify a 133 bp PCR product from the multi-copy, highly conserved 18S ribosomal RNA genes of Pf. qPCR used the following adapted oligonucleotide primers and probe^9^: 18s forward primer 5’-GTA ATT GGA ATG ATA GGA ATT TAC AAG GT-3’, 18s reverse primer 5’-TCA ACT ACG AAC GTT TTA ACT GCA AC-3’, and FAM labelled probe sequence 5’-FAM- AAC AAT TGG AGG GCA AG-NFQ-MGB-3’, with TaqMan Universal PCR master mix II (with Amperase UNG), and was run on an QuantStudio3 machine with DA2 software (Applied Biosystems). Default Universal qPCR (target FAM-NFQ-MGB) and quality control (QC) settings were used. The standard curve utilized known quantities of cultured Pf material, as previously described^6^.

For QC purposes, qPCR samples were re-tested if replicates included a mixture of positive and negative (in terms of amplification) results with one or more positive results >100 gc/mL or if the % CV of any results were high outliers. All ‘passed’ data following the quality control steps above, including any 0 values, were used to generate the final mean qPCR result for each time-point.

#### Thick blood film microscopy

Blood film microscopy was used in phases VAC069A and VAC069B only. Collection of blood, preparation of thick films and slide reading were performed according to Jenner Institute Standard Operating Procedure (SOP) ML009. Slides were prepared using Field’s stain A and then Field’s stain B. 200 fields at high power (1000x) were read. Visualization of two or more parasites in 200 high power fields constituted a positive result. For internal quality control, all slides were read separately by two microscopists, with a third read if results were discordant (one negative and one positive report).

#### Diagnostic algorithms used in VAC069 study

Algorithm for diagnosis and initiation of treatment VAC069A and B:
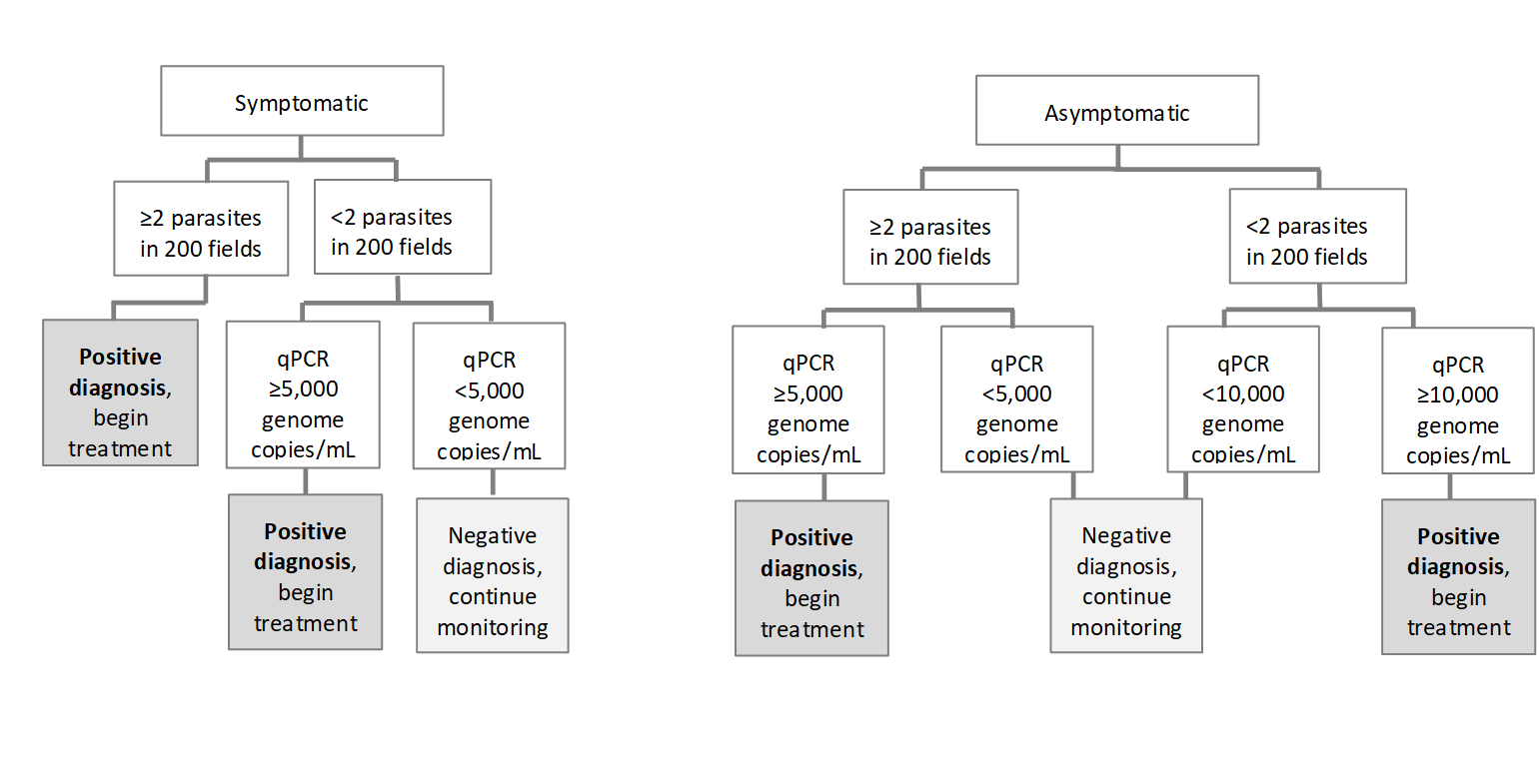

Algorithm for diagnosis and initiation of treatment in VAC069C to E:
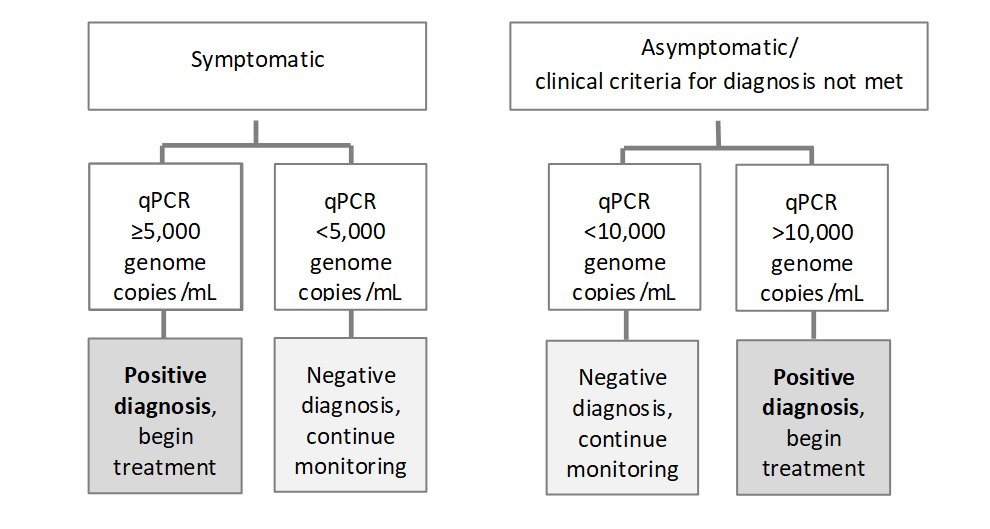

#### Modelling of parasite multiplication rate

A qPCR-derived parasite multiplication rate (PMR) was modelled based on previously described methodology with modifications ^5,8^. The arithmetic mean of three replicate qPCR results obtained for each individual at each time-point was used for model-fitting. Negative individual replicates were assigned a value of 0 gc/mL for the purposes of calculating the arithmetic mean of triplicates (where at least one of the three readings was positive). All qPCR data points which, based upon the mean of the three replicates, were greater than 5 gc/mL were used for modelling. Any values ranging from 1-5 gc/mL and any data point that was negative but preceded a positive data point were replaced with a value of 5 gc/mL; otherwise negative data points occurring after any positive data point but not preceding a positive data point were treated as 0 gc/mL. The time interval between the morning and evening bleeds used for qPCR monitoring was set as 0.37 days. PMR per 48 h was then calculated using a linear model fitted to log_10_-transformed qPCR data.

To allow direct comparison across the different cohorts for *P. vivax* CHMI, data from timepoints in CHMIs conducted in September 2019 and May 2021, which would not have been available if using the visit schedule for the *P. vivax* CHMI in October 2021, were removed prior to model-fitting.

### Supplementary Tables

#### Table S1: Demographics of study participants.

*Pv* CHMI-1 indicates participants who underwent primary controlled human malaria infection with *P. vivax*; *Pv* CHMI-2 secondary homologous *P. vivax* CHMI; *Pv* CHMI-3 tertiary homologous *P. vivax* CHMI; *Pf* CHMI repeat heterologous CHMI with *P. falciparum*.

|  | | **VAC069** | | | | **VAC079** | |
| --- | --- | --- | --- | --- | --- | --- | --- |
|  |  | ***Pv* CHMI-1** | ***Pv* CHMI-2** | ***Pv* CHMI-3** | ***Pf* CHMI** | ***Pv* CHMI-1** | ***Pv* CHMI-2** |
| **No. of participants** | | 19 | 12 | 2 | 6 | 10 | 5 |
| **Female Sex, no. (%)** | | 9 (47.4) | 6 (50.0) | 0 | 4 (66.7) | 8 (80.0) | 5 (100.0) |
| **Age**  **(yr)** | **Median** | 26 | 27 | 30 | 33 | 37 | 42 |
|  | **Range** | 21-48 | 22-48 | 29-31 | 22-49 | 22-45 | 23-46 |
| **Body Mass Index (kg/m^2^)** | **Median** | 23.6 | 24.1 | 28.7 | 25.1 | 28.9 | 27.6 |
|  | **Range** | 17.5-33.5 | 18.1-32.6 | 24.8-32.6 | 18.9-33.5 | 23.5-40.2 | 23.5-35.9 |
| **Ethnicity,**  **no. (%)** | **White-British** | 8 (42.1) | 5 (41.7) | 1 (50.0) | 4 (66.7) | 9 (90.0) | 4 (80.0) |
|  | **White-Other** | 6 (31.6) | 4 (33.3) | 1 (50.0) | 2 (33.3) | 1 (10.0) | 1 (20.0) |
|  | **Asian** | 2 (10.5) | 1 (8.33) | 0 | 0 | 0 | 0 |
|  | **Arab** | 2 (10.5) | 1 (8.33) | 0 | 0 | 0 | 0 |
|  | **Mixed** | 1 (5.3) | 1 (8.33) | 0 | 0 | 0 | 0 |
| **Duffy Phenotype,**  **no. (%)** | **Fy(a+b-)** | 5 (26.3) | 4 (33.3) | 1 (50.0) | 0 | 1 (10.0) | 0 |
|  | **Fy(a-b+)** | 3 (10.5) | 2 (16.7) | 0 | 1 (16.7) | 3 (30.0) | 2 (40.0) |
|  | **Fy(a+b+)** | 11 (63.2) | 6 (50.0) | 1 (50.0) | 5 (83.3) | 6 (60.0) | 3 (60.0) |

#### Table S2: Unsolicited adverse events.

Unsolicited adverse events (AEs) deemed possibly, probably or definitely related to CHMI in each study. Number of episodes of unsolicited AE are listed by MEDDRA System Organ Class and Preferred Term and maximal severity grade.

**Table S2A Unsolicited AEs in the VAC069 study during *P. vivax* CHMI**

| **Unsolicited adverse event** | | **Primary CHMI** (n=19) | | **Secondary CHMI** (n=12) | | **Tertiary CHMI** (n=2) | | **Total** |
| --- | --- | --- | --- | --- | --- | --- | --- | --- |
| System Order Class | Preferred Term | Grade 1 | Grade 2 | Grade 1 | Grade 2 | Grade 1 | Grade 2 |  |
| Blood and lymphatic system disorders | Lymphadenopathy | 1 |  |  |  |  |  | 1 |
| Gastrointestinal disorders | Constipation | 1 |  |  |  |  |  | 1 |
|  | Tooth infection |  |  |  | 1 |  |  | 1 |
|  | Dyspepsia |  |  |  |  | 1 |  | 1 |
| Infections and infestations | Tonsillitis |  |  |  | 1 |  |  | 1 |
| Metabolism and nutrition disorders | Decreased appetite | 2 | 1 |  |  |  |  | 3 |
| Musculoskeletal and connective tissue disorders | Myalgia | 1 |  |  |  |  |  | 1 |
|  | Musculoskeletal stiffness | 1 |  |  |  |  |  | 1 |
| Nervous system disorders | Paraesthesia | 1 |  |  |  |  |  | 1 |
| Psychiatric disorders | Stress | 1 |  |  |  |  |  | 1 |
| Respiratory, thoracic and mediastinal disorders | Dry throat | 1 |  |  |  |  |  | 1 |
|  | Oropharyngeal pain | 1 |  |  |  |  |  | 1 |
| Skin and subcutaneous tissue disorders | Pruritus | 1 |  |  |  |  |  | 1 |

**Table S2B Unsolicited AEs in the VAC079 study during *P. vivax* CHMI**

| **Unsolicited adverse event** | | **Primary CHMI** (n=10) | | **Secondary CHMI** (n=5) | | **Total** |
| --- | --- | --- | --- | --- | --- | --- |
| System Order Class | Preferred Term | Grade 1 | Grade 2 | Grade 1 | Grade 2 |  |
| Gastrointestinal disorders | Abdominal pain upper |  |  |  | 1 | 1 |
| General disorders and administration site conditions | Chest pain |  |  | 1 |  | 1 |
| Metabolism and nutrition disorders | Decreased appetite | 1 |  |  |  | 1 |
| Musculoskeletal and connective tissue disorders | Musculoskeletal stiffness | 1 |  |  |  | 1 |
|  | Back pain | 1 |  |  |  | 1 |
| Nervous system disorders | Paraesthesia | 1 |  |  |  | 1 |
| Renal and urinary disorders | Chromaturia | 2 |  |  |  | 2 |
| Skin and subcutaneous tissue disorders | Rash | 1 |  |  |  | 1 |

**Table S2C Unsolicited AEs in the VAC069 study during *P. falciparum* CHMI**

| **Unsolicited adverse event** | | ***P. falciparum* CHMI** (n=6) | **Total** |
| --- | --- | --- | --- |
| System Order Class | Preferred Term | Grade 1 |  |
| Infections and infestations | Nasopharyngitis | 1 | 1 |
| Skin and subcutaneous tissue disorders | Rash maculo-papular | 1 | 1 |

#### Table S3: Laboratory adverse events.

Laboratory adverse events (AEs) deemed at least possibly related to CHMI in each study. Number of episodes of AE and maximal severity of AE during episode listed.

**Table S3A Laboratory AEs in the VAC069 study during *P. vivax* CHMI**

| **Laboratory abnormality** | | **Primary CHMI** (n=19) | | | **Secondary CHMI** (n=12) | | | **Tertiary CHMI** (n=2) | | | **Total** |
| --- | --- | --- | --- | --- | --- | --- | --- | --- | --- | --- | --- |
|  |  | Grade 1 | Grade 2 | Grade 3 | Grade 1 | Grade 2 | Grade 3 | Grade 1 | Grade 2 | Grade 3 |  |
| Haematology | Leucopaenia | 9 | 5 |  | 1 |  |  |  |  |  | 15 |
|  | Lymphopaenia | 5 | 7 | 5 | 1 | 1 |  | 1 |  |  | 20 |
|  | Neutropaenia | 5 | 1 |  | 1 |  |  | 1 |  |  | 8 |
|  | Anaemia | 4 | 2 |  | 5 | 1 |  |  |  |  | 12 |
|  | Thrombocytopaenia | 3 | 4 | 2 | 1 |  |  |  |  |  | 10 |
| Biochemistry | Raised ALT | 5 | 4 | 3 | 1 |  |  |  | 1 |  | 14 |
|  | Hyperbilirubinaemia | 1 |  |  |  |  |  |  |  |  | 1 |
|  | Hypoalbuminaemia |  |  |  | 1 |  |  |  |  |  | 1 |
|  | Raised urea |  |  |  | 1 |  |  |  |  |  | 1 |
|  | Hypokalaemia | 2 |  |  | 1 |  |  | 1 |  |  | 4 |

**Table S3B Laboratory AEs in the VAC079 study during *P. vivax* CHMI**

| **Laboratory abnormality** | | **Primary CHMI** (n=10) | | | **Secondary CHMI** (n=5) | | | **Total** |
| --- | --- | --- | --- | --- | --- | --- | --- | --- |
|  |  | Grade 1 | Grade 2 | Grade 3 | Grade 1 | Grade 2 | Grade 3 |  |
| Haematology | Leucoytosis |  |  |  | 1 |  |  | 1 |
|  | Leucopaenia | 1 | 4 |  |  |  |  | 5 |
|  | Lymphopaenia |  | 5 | 2 |  |  |  | 7 |
|  | Neutropaenia | 3 | 1 |  |  |  |  | 4 |
|  | Anaemia | 5 |  |  | 3 |  |  | 8 |
|  | Thrombocytopaenia |  |  | 1 |  |  |  | 1 |
| Biochemistry | Raised ALT | 1 | 5 | 1 |  |  |  | 7 |
|  | Raised ALP | 2 |  |  |  |  |  | 2 |

**Table S3C Laboratory AEs in the VAC069 study during *P. falciparum* CHMI**

| **Laboratory abnormality** | | ***P. falciparum* CHMI** (n=6) | | | **Total** |
| --- | --- | --- | --- | --- | --- |
|  |  | Grade 1 | Grade 2 | Grade 3 |  |
| Haematology | Leucopaenia | 2 | 1 |  | 3 |
|  | Lymphopaenia |  | 1 | 3 | 4 |
|  | Neutropaenia | 3 |  |  | 3 |
|  | Anaemia | 1 |  |  | 1 |
|  | Thrombocytopaenia | 1 | 1 |  | 2 |
| Biochemistry | Raised creatinine | 1 |  |  | 1 |
|  | Raised ALT | 2 |  | 1 | 3 |
|  | Hyperbilirubinaemia | 1 |  |  | 1 |

#### Table S4: Malaria qPCR data (gc/mL) for each CHMI

Malaria qPCR data used in PMR modelling are shown. The top row represents day (D) of follow-up visit post blood-stage CHMI. DoD indicates the timepoint at which malaria diagnostic criteria were reached. Treatment in some participants was started half a day after reaching malaria diagnostic criteria. qPCR data shown for samples taken prior to starting treatment. qPCR negative values for all three triplicate readings in the assay are indicated by ‘N’. Squares highlighted in gray indicate negative or < 20 gc/mL which is below minimum positive reporting criteria and these datapoints were removed for PMR modelling. Datapoints which would not have been taken during CHMI in October 2021 due to changes in protocol were removed for PMR modelling and are not shown. Blacked out boxes indicate the timepoints after a participant commenced antimalarial treatment.

**Table S4A Malaria qPCR during VAC069A in Jan 2019**

| **No of CHMI** | **DoD** | **D7** | **D8** | **D9** | **D10** | **D10.5** | **D11** | **D11.5** | **D12** | **D12.5** | **D13** | **D13.5** | **D14** | **D14.5** | **D15** | **D15.5** | **D16** | **D16.5** | **D17** |
| --- | --- | --- | --- | --- | --- | --- | --- | --- | --- | --- | --- | --- | --- | --- | --- | --- | --- | --- | --- |
| 1 (1 vial) | 15.5 | 17 | 17 | 58 | 67 |  | 301 |  | 294 |  | 956 |  | 2251 | 417 | 5035 | 3779 |  |  |  |
| 1 (1 vial) | 12.5 | 78 | 60 | 508 | 1060 | 2060 | 3132 | 2640 | 5393 | 11259 |  |  |  |  |  |  |  |  |  |
| 1 (1:5) | 15 | 5 | 18 | 72 | 239 |  | 519 |  | 972 |  | 2666 | 2999 | 4700 | 1538 | 17795 | 14843 |  |  |  |
| 1 (1:5) | 15 | 6 | 21 | 33 | 8 |  | 266 |  | 459 |  | 1331 |  | 2632 | 1056 | 8687 | 7112 |  |  |  |
| 1 (1:20) | 15.5 | N | 5 | 32 | 57 |  | 406 |  | 616 |  | 1431 | 1253 | 3054 | 1444 | 9597 | 8134 | 15768 |  |  |
| 1 (1:20) | 16.5 | N | 5 | 15 | 8 |  | 53 |  | 76 |  | 239 |  | 717 |  | 1479 | 1438 | 2593 | 9668 | 7894 |

**Table S4B Malaria qPCR during VAC069B in Sept 2019**

| **No of CHMI** | **DoD** | **D7** | **D8** | **D9** | **D10** | **D10.5** | **D11** | **D11.5** | **D12** | **D12.5** | **D13** | **D13.5** | **D14** | **D14.5** | **D15** | **D15.5** | **D16** | **D16.5** | **D17** |
| --- | --- | --- | --- | --- | --- | --- | --- | --- | --- | --- | --- | --- | --- | --- | --- | --- | --- | --- | --- |
| 1 | 15.5 | N | 5 | 45 | 104 |  | 209 |  | 270 |  | 1379 | 1391 | 2244 | 3670 | 8921 | 9574 | 16345 |  |  |
| 1 | 14.5 | 5 | 9 | 72 |  | 5 | 265 |  | 778 |  | 3365 | 2835 | 4283 | 17392 | 19589 |  |  |  |  |
| 2 | 21 | N | N | N | N | N | N | 5 | 32 |  | 70 |  | 119 |  | 235 |  | 217 |  | 995 |
| 2 | 15.5 | 5 | 20 | 41 | 17 |  | 113 |  | 351 |  | 1371 | 492 | 2299 | 3374 | 8679 | 8489 | 17449 |  |  |
| 2 | 14 | 11 | 68 | 86 | 145 |  | 553 |  | 1405 | 2601 | 5691 | 4021 | 11364 | 17496 |  |  |  |  |  |

| **No of CHMI** | **DoD** | **D17.5** | **D18** | **D18.5** | **D19** | **D19.5** | **D20** | **D20.5** | **D21** | **D21.5** |
| --- | --- | --- | --- | --- | --- | --- | --- | --- | --- | --- |
| 1 | 15.5 |  |  |  |  |  |  |  |  |  |
| 1 | 14.5 |  |  |  |  |  |  |  |  |  |
| 2 | 21 |  | 1180 | 1033 | 3119 | 3026 | 3950 | 2713 | 11194 | 12522 |
| 2 | 15.5 |  |  |  |  |  |  |  |  |  |
| 2 | 14 |  |  |  |  |  |  |  |  |  |

**Table S4C Malaria qPCR during VAC069C in May 2021**

| **No of CHMI** | **DoD** | **D7** | **D8** | **D9** | **D10** | **D11** | **D11.5** | **D12** | **D12.5** | **D13** | **D13.5** | **D14** | **D14.5** | **D15** | **D15.5** | **D16** | **D16.5** | **D17** | **D17.5** | **D18** | **D18.5** | **D19** |
| --- | --- | --- | --- | --- | --- | --- | --- | --- | --- | --- | --- | --- | --- | --- | --- | --- | --- | --- | --- | --- | --- | --- |
| 1 | 15.5 | N | 5 | 74 | 220 | 316 |  | 408 |  | 1678 | 1315 | 3625 | 6183 | 8674 | 9095 |  |  |  |  |  |  |  |
| 1 | 18 | 10 | 5 | 8 | N | 125 |  | 88 |  | 291 |  | 814 | 1252 | 1652 | 1805 | 2152 | 6550 | 7261 | 6161 | 12050 | 20348 |  |
| 1 | 19 | N | 5 | 26 | 37 | 98 |  | 62 |  | 268 |  | 298 |  | 1111 | 1262 | 1617 | 2121 | 6005 | 6395 | 4758 | 9432 | 19781 |
| 1 | 14 | 32 | 35 | 108 | 182 | 1059 | 748 | 1121 | 2572 | 8025 | 7805 | 14766 | 18983 |  |  |  |  |  |  |  |  |  |
| 1 | 16 | 11 | 5 | 47 | 74 | 311 |  | 298 |  | 1669 | 1544 | 3316 | 6509 | 8709 | 9292 | 15568 | 39496 |  |  |  |  |  |
| 1 | 14.5 | N | 5 | 50 | 96 | 370 |  | 754 |  | 2641 | 2335 | 4847 | 12160 | 7238 |  |  |  |  |  |  |  |  |
| 1 | 15 | N | 5 | 113 | 183 | 485 |  | 556 |  | 3001 | 2476 | 5926 | 7904 | 16068 | 20776 |  |  |  |  |  |  |  |
| 2 | 15 | 5 | 35 | 70 | 93 | 538 |  | 870 |  | 3906 | 3531 | 7560 | 9434 | 22138 | 24051 |  |  |  |  |  |  |  |
| 2 | 15 | 25 | 54 | 82 | 103 | 447 |  | 450 |  | 2626 | 2679 | 4126 | 6192 | 13673 | 14529 |  |  |  |  |  |  |  |
| 3 | 14.5 | 16 | 9 | 92 | 122 | 331 |  | 480 |  | 2780 | 2934 | 6908 | 13567 | 22850 |  |  |  |  |  |  |  |  |

**Table S4D Malaria qPCR during VAC069D in Oct 2021**

| **No of CHMI** | **DoD** | **D7** | **D8** | **D9** | **D10** | **D10.5** | **D11** | **D11.5** | **D12** | **D12.5** | **D13** | **D13.5** | **D14** | **D14.5** | **D15** | **D15.5** | **D16** | **D16.5** | **D17** | **D17.5** | **D18** |
| --- | --- | --- | --- | --- | --- | --- | --- | --- | --- | --- | --- | --- | --- | --- | --- | --- | --- | --- | --- | --- | --- |
| 1 | 15 | 8 | 28 | 66 | 124 |  | 604 |  | 806 |  | 2823 | 2487 | 3759 | 3551 | 14373 | 10759 |  |  |  |  |  |
| 1 | 15 | 17 | 17 | 71 | 119 |  | 1021 | 804 | 1617 | 1727 | 3405 | 4145 | 5742 | 6828 | 17155 | 13910 |  |  |  |  |  |
| 1 | 17 | 5 | 9 | 5 | 7 |  | 73 |  | 46 |  | 510 |  | 501 |  | 1735 | 1792 | 3600 | 4258 | 12302 | 13366 |  |
| 1 | 15.5 | 5 | 61 | 53 | 73 |  | 401 |  | 767 |  | 1552 | 1096 | 2597 | 2245 | 5737 | 3057 |  |  |  |  |  |
| 2 | 16 | 16 | 16 | 92 | 24 |  | 504 |  | 393 |  | 1536 | 1595 | 1702 | 3079 | 7646 | 6541 | 12156 | 16931 |  |  |  |
| 2 | 16.5 | 9 | 5 | 31 | 84 |  | 195 |  | 303 |  | 998 |  | 1180 | 2409 | 3554 | 3209 | 6561 | 13796 | 16362 |  |  |
| 2 | 17.5 | 5 | 19 | 77 | 58 |  | 384 |  | 310 |  | 565 |  | 1056 | 839 | 2261 | 1844 | 3977 | 6472 | 9923 | 12001 | 16503 |
| 2 | 14 | 8 | 60 | 200 | 271 |  | 1631 | 996 | 2660 | 5989 | 8523 | 8670 | 12188 | 36683 |  |  |  |  |  |  |  |
| 2 | 15 | 17 | 18 | 86 | 139 |  | 732 |  | 861 |  | 2866 | 2681 | 3215 | 4970 | 12803 | 9949 |  |  |  |  |  |
| 2 | 14.5 | 5 | 59 | 56 | 205 |  | 651 |  | 1436 | 1627 | 3081 | 2283 | 5242 | 10170 | 16545 |  |  |  |  |  |  |
| 2 | 15 | 9 | 69 | 87 | 138 |  | 735 |  | 1096 | 1349 | 3599 | 3153 | 6065 | 9292 | 23263 | 22349 |  |  |  |  |  |
| 3 | 14 | 5 | 38 | 232 | 213 |  | 1447 | 1516 | 2393 | 3266 | 7133 | 8062 | 12560 | 19858 |  |  |  |  |  |  |  |

**Table S4E Malaria qPCR during VAC069E in Oct 2022**

| **No of CHMI** | **DoD** | **D5** | **D6** | **D7** | **D7.5** | **D8** | **D8.5** | **D9** | **D9.5** | **D10** | **D10.5** | **D11** | **D11.5** | **D12** |
| --- | --- | --- | --- | --- | --- | --- | --- | --- | --- | --- | --- | --- | --- | --- |
| 3 | C + 10 | N | N | 157 | 83 | 171 | 570 | 1752 | 313 | 16701 | 11670 |  |  |  |
| 3 | C + 9 | 21 | 10 | 518 | 326 | 1557 | 3247 | 10161 |  |  |  |  |  |  |
| 3 | C + 8.5 | N | 19 | 965 | 741 | 3780 | 11311 |  |  |  |  |  |  |  |
| 2 | C + 10 | 4 | N | 139 | 39 | 694 | 1060 | 1951 | 953 | 11790 | 24274 |  |  |  |
| 2 | C + 10 | N | N | 164 | 153 | 427 | 1027 | 3116 | 1472 | 33908 | 32777 |  |  |  |
| 2 | C + 12 | 4 | 5 | 30 |  | 178 |  | 695 |  | 9563 | 4890 | 1854 | 7059 | 127550 |

#### Table S5: Merozoite antigens tested in multiplex immunoassay

| Gene ID | Addgene ID | Name | Description | Domain Boundary | CD4 tag (20.4kDa)  Present | Mol.wt. (kDa)  Including Tag | Bead Region | Working stock concentrationml^-1^ |
| --- | --- | --- | --- | --- | --- | --- | --- | --- |
| KMZ83376.1 | - | PvEBP | Plasmodium vivax  Erythrocyte Binding  Protein | M1-716P | No | 95 | 12 | 0.7x10^6^ |
| PVX_113775 | #68516 | Pv12 | Plasmodium vivax  6-cysteine protein | F24-A339 | No | 38.6 | 14 | 0.72x10^6^ |
| PVP01_0934200 | - | PvAMA1 | Plasmodium vivax  Apical Merozoite Antigen 1 | K25-479E | No | 52.7 | 15 | 0.74x10^6^ |
| PVX_099980 | #68505 | PvMSP1 | Plasmodium vivax  Merozoite Surface Protein 1 | E20-P1721 | No | 215 | 18 | 0.59x10^6^ |
| PVX_088910 | #68522 | PvGAMA | Plasmodium vivax  GPI-anchored micronemal antigen, putative | L21-S749 | No | 85 | 21 | 1.32x10^6^ |
| PVP01_0000100 | - | PvTRAg25 | Plasmodium vivax Tryptophan Rich Antigen 25 | K62-L693 | No | 74.3 | 22 | 0.75x10^6^ |
| PVX_110810 | #68528 | PvDBP | Plasmodium vivax Duffy binding protein | V23-T1008 | Yes | 142 | 25 | 0.9x10^6^ |
| Cd4 | - | CD4 | Cluster of differentiation 4 | S1-183N | - | 20.4 | 20 | 0.5x10^6^ |
